# A Generative Patient Digital Twin for Sequential Treatment of Oropharyngeal Squamous Carcinomas

**DOI:** 10.1101/2025.10.10.25337766

**Authors:** Zishun Yu, Harshal Jagdishbhai Hirpara, Xinhua Zhang, Guadalupe Canahuate, Andrew Wentzel, Yaohua Wang, Carla Floricel, Serageldin K. Attia, Abdallah SR Mohamed, Mohamed Naser, Clifton D Fuller, G Elisabeta Marai

**Affiliations:** Department of Computer Science, University of Illinois at Chicago, Chicago, IL, United States; Department of Electrical and Computer Engineering, University Of Iowa, Iowa City, IA, United States; Department of Radiation Oncology, The University of Texas MD Anderson Cancer Center, Austin, TX, United States

## Abstract

Real-world testing of treatment strategies is often infeasible, emphasizing the need for robust simulation frameworks that can model diverse patient characteristics and predict treatment outcomes. In this study, we present a generative simulator designed to synthesize patient profiles and forecast treatment results under hypothetical scenarios,with the goal of facilitating personalized treatment planning with what-if scenarios. The proposed simulator is able to predict disease progression and treatment outcomes based on synthesized profiles through application of Variational Autoencoder and XGBoost models. Simulations evaluated its ability of generating realistic baseline patient profiles, and the predictive accuracy of the combined framework. The proposed method outperformed rule-based approaches and multilayer perceptron models in predicting 22 out of 25 clinical variables, with performance measured by F1 scores for categorical variables and Mean Squared Error for numerical variables. Case studies of two patients drawn from ground truth data illustrate that the simulator framework can represent both short treatment courses with early relapse and prolonged multi-modal trajectories with recurrent disease. These results underscore the framework’s ability to capture complex relationships in clinical data and highlight its advantages over baseline methods. Although this work focuses on validating the simulator’s generative and predictive capabilities, it establishes a foundation for future research in personalized treatment planning, including what-if analyses and reinforcement learning studies.

## Introduction

Head and neck cancer, which includes cancers of the larynx, throat, lips, mouth, nose, and salivary glands, is now an epidemic, with 65,000 new cases in the United States annually [1], whose treatment is, as in many other types of cancers, a dynamic and complex process. This therapy process involves making multiple, patient-specific treatment decisions to maximize efficacy—for example, reduction in tumor size, time of local region control, and survival time—while minimizing side effects [2-4]. For example, a specific patient may undergo definitive surgery (DS), induction therapy (IC), concurrent therapy (CC), and/or radiation therapy (RT) [5]. After each round of IC, a decision must be made whether to continue IC or start either RT or CC. These decisions are currently taken by clinicians or multidisciplinary tumor boards based on pretherapy patient characteristics or crude heuristics. Notably, current risk-prediction models (eg, American Joint Committee on Cancer [AJCC] staging) incorporated in clinical decision support systems do not by themselves systematically direct clinicians to select an appropriate treatment that incorporates both oncologic and toxicity endpoints. Furthermore, disposition to initial IC is then followed by a second responsive disposition to either RT or concurrent chemoradiotherapy. Inferring the optimal treatment policies for multistage decisions (eg, which treatment to administer initially and then after observing treatment response; Figure 1) post hoc is challenging because an optimal therapy sequence cannot be readily pieced together from several single-stage decisions. For this reason, in the absence of rigorous clinical trials comparing adaptive IC permutations with concurrent RT, group comparison is exceedingly difficult because simple models that account for confounders at initial disposition (eg, propensity scores) are unequipped to incorporate sequential decision processes (eg, the choice of CC after IC).

**Figure 1.**
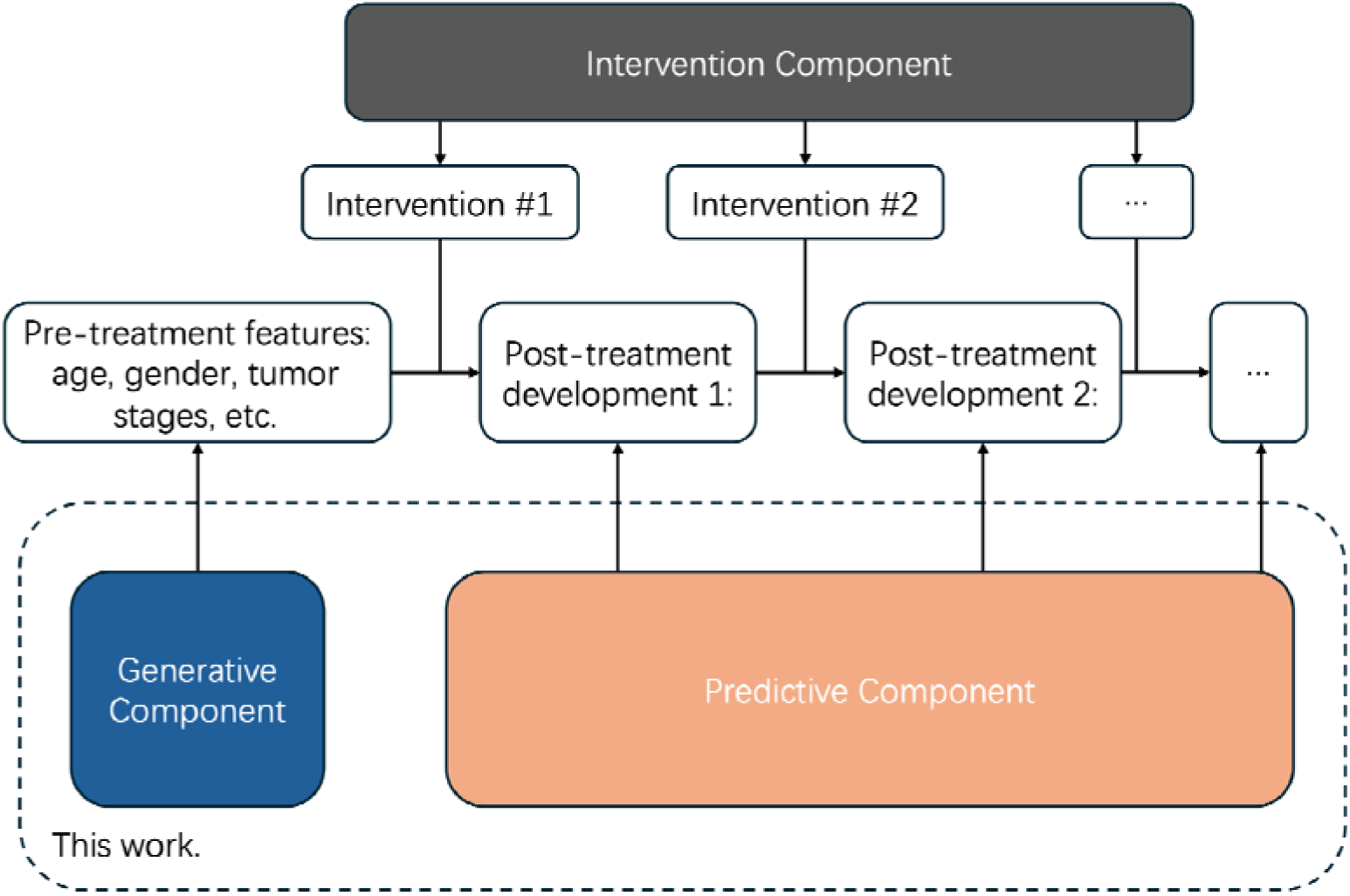
An overview of our digital twin design

Digital twins are a concept used in industry, where a digital replica of a physical entity or process is virtually recreated, with similar elements and dynamics, to perform real-time optimization and testing [6]. This concept has been adapted to health care where machine learning models have been trained to allow personalization and optimization of health outcomes, prediction and prevention of adverse events, and planning interventions [7, 34].

Several examples highlight the application of digital twins in healthcare. For sepsis patients, digital twins have been developed to simulate patient conditions and predict adverse events in real-time, aiding in timely interventions [8]. In stroke patients, digital twin models are being used to predict recovery trajectories and personalize rehabilitation plans to optimize care [9]. Similarly, digital twins have been applied to model vascular blood flow, enhancing the understanding of cardiovascular conditions and informing treatment strategies [10]. For lung cancer, digital twins are used to optimize radiotherapy treatments by simulating how tumors and surrounding tissues will respond to different treatment plans [11]. A recent study demonstrates the use of digital twins to support multiple stages of personalized cancer treatment, showcasing their ability to enhance clinical decision-making and treatment optimization in oncology [12]. A broader survey of current methods outlines the diverse applications of digital twins in healthcare, illustrating their potential across various stages of the patient journey [13, 14]. The most relevant work is perhaps [33], where the authors design a digital twin for head and neck cancer. The key difference between their work and ours is that their approach is *predictive*, while ours is *generative*. Specifically, their work uses a test set of patients and employs support vector machines (SVMs) to predict disease progression for treatment plans suggested by the model. In contrast, our approach first fits a patient generator that aligns its distribution with real patient data to generate new synthetic patients, rather than directly evaluating on test data. We then predict disease progression using XGBoost. The generative nature of our simulator offers several advantages: (1) it enables the creation of diverse synthetic patient scenarios, allowing exploration of rare or unobserved cases while reducing the risk of overfitting to the test data; (2) by simulating a broader patient population, it incorporates stochastic variability in disease progression, improving the robustness and generalizability of predictive insights.

**Potential uses** for patient digital twins can help generate synthetic data for training healthcare models, exposing downstream safety issues by exploring dangerous scenarios, and predicting and visualizing [15] potential outcomes for physicians. Additionally, digital twins can help support what-if analysis, where alternative treatments or conditions can be explored by physicians, and counterfactual generation, where a minimal set of changes can be identified to create a different outcome for each patient. These technologies can be used to investigate different scenarios and tailor treatments to individual patients. Digital twins are also used as building blocks for sequential treatment selection models such as Reinforcement Learning (RL) models. Directly applying AI-generated treatment to human subjects is generally unethical, differing from many other RL applications such as game playing (e.g., AlphaGo). This necessitates offline RL, which relies on online finetuning to perform competitively. Therefore, interaction with the environment is required. Furthermore, although off-policy evaluation of RL algorithms has been well studied [6], they generally suffer high variance, and it is more straightforward if a digital twin of cancer treatment is available, allowing us to simulate the evolution of a patient’s condition under the AI-proposed treatment history, hence evaluate it.

A well calibrated and robust digital twin is necessary before it is feasible to implement any other downstream task. Therefore, in this work we introduce an approach based on digital twinning to model head and neck cancer patients’ response to treatment, survival and toxicity outcomes. By leveraging a large number of head and neck cancer cases collected at a single institutional head and neck data tumor board at the MD Anderson Cancer Center (MDACC), we propose a methodology approach to leverage generative models and XGBoost models to construct a digital twin dyad for simulation of therapy outcomes.

## Methods

### Overview

An ideal **digital twin** for healthcare should contain two major components: (1) a **generative** [17-19] component: it generates a new patient’s initial status before any treatment intervention so that one could study how different interventions affect the subsequent disease development. The generative component should fit the patient status and symptom distribution to accurately represent the population; (2) a **prediction** [20-22] component: based on the received/generated patient status, the predictive component should accurately forecast the development of disease for different interventions, to enhance the understanding of different clinical actions.

To this end, we design our digital twin simulator as follows: (1) using a Variational Autoencoder (VAE) [17] as the generative component; and (2) XGBoost [20] as the predictive component to forecast the future development of disease.

### Variational auto-encoder

A Variational Autoencoder (VAE) is a generative model designed to learn a compressed representation of high-dimensional data, such as medical images [23] or patient data [24], while preserving essential patterns and structures. VAEs utilize probabilistic inference [25], enabling them to approximate complex data distributions and generate new samples that are similar to the original dataset. This ability makes them particularly useful in medical research for tasks such as disease modeling [23-24], anomaly detection [26], and synthesizing medical images [23]. By encoding data into a lower-dimensional latent space [27] and then reconstructing it, VAEs can capture nuanced relationships in complex medical data, offering insights into underlying trends or rare conditions.

### XGBoost

XGBoost is a gradient-boosting [28] algorithm widely used for its efficiency and predictive accuracy in large-scale datasets. It operates by constructing an ensemble of decision trees [29] in a sequential manner, with each new tree correcting the errors of the previous ones. In the medical domain, XGBoost has been applied successfully to various predictive modeling tasks, such as identifying disease risk factors [30], predicting patient outcomes [31], and classifying clinical conditions [32] based on diverse data types (e.g., imaging, lab results, and electronic health records). Its scalability, combined with its ability to handle missing data and reduce overfitting, makes it a robust tool for handling complex medical datasets with high dimensionality.

## Data Descriptions

### Data overview

We performed a retrospective review of 676 patients with oropharyngeal squamous cell carcinoma who were treated using radiation therapy (RT) at the MDACC between 2010 and 2021. The patient data, extracted from medical records, include clinical and treatment information, and patient-reported symptom ratings. The *clinical* attributes used in this work include demographics: age, gender, and smoking status. The *diagnostic* attributes include tumor size, lymph node stage, and tumor sub-site. The *treatment* attributes include indicators as to whether the patient received definitive surgery (DS), induction therapy (IC), concurrent therapy (CC), and/or radiation therapy (RT).

The 676 patient examples were partitioned into two disjoint sets for training and testing, using a 80% (n=539)-20% (n=137) random split.

Each patient is associated with a sequence of features, depending on the duration and frequency of follow-up assessments. These features capture a wide range of clinical and patient-reported data, reflecting different aspects of the treatment and recovery process. Given the diverse nature of these features, we have organized them into five distinct categories for the design of our simulator. It is important to emphasize that these categories are not intended to represent a strict temporal progression, as they can occur at different time points in the patient’s treatment process. In the Temporal Modeling section, we will provide a detailed visualization of how the temporal dependencies between these feature categories unfold.

- **Pre-treatment features** (PTF): These are baseline characteristics measured before the initiation of any clinical interventions. Examples include demographic and lifestyle factors such as age, gender, smoking status, and other relevant patient history. These features are essential for understanding the patient’s initial health state and predicting how they might respond to treatment.
- **Interventions/actions** (Actions): This category encompasses the medical treatments or procedures undertaken to address the patient’s condition. It includes both surgical and non-surgical interventions such as surgery, chemotherapy, radiotherapy, and other therapies. These actions are a critical component of the treatment process, directly influencing patient outcomes.
- **Post-intervention outcomes** (PIO): These refer to the measurable outcomes that occur following clinical interventions. This could include variables such as the specific type of radiotherapy administered, the dosage of radiotherapy, and the immediate effects observed as a result of the intervention. Post-intervention outcomes help in assessing the efficacy and safety of the treatments. Technically, dosage is a parameter of radiotherapy (an action), but we treat it as an outcome in the sense of “now that we’ve decided to carry out radiotherapy, the dosage can be worked out from a standard procedure”. This indeed makes our simulator more useful, tasking it to additionally simulate the dosage instead of relying on the user’s specifications.
- **Patient report scores** (PRS): Symptom burden data are collected using patient-reported scores (PRS) questionnaires based on MDASI-HN (MD Anderson Symptom Inventory, the Head and Neck Module) [16], a 28 symptom inventory. In the questionnaire, patients are asked to rate symptoms using a 0-to-10 scale, from ‘‘not present’’ (0) to ‘‘as bad as you can imagine’’ (10). Symptoms are grouped into 3 categories: HNC-specific, general cancer, and six interference symptoms. In this work, we focus on the 4 symptoms (fatigue, drymouth, swallow and taste) as examples due to space constraint, listed as a reference in Table 4. PRS data are collected prior to treatment and subsequently at multiple points during and after the treatment process. During treatment, a spike in symptom burden is expected due to treatment toxicity with most symptoms subsiding over time. However, for some patients, toxicity treatment leads to long-lasting sequelae. Dry mouth and taste are some of the most prevalent symptoms for oropharyngeal cancer patients.
- **Treatment plan outcomes** (TPO): This category reflects the overall success or failure of the entire treatment plan. It evaluates the long-term effectiveness of the combined interventions and provides an integrated measure of the patient’s health trajectory, including whether the treatment achieved its intended goals.

Table 1, Table 2, and Table 3 collectively provide an overview of the data characteristics in our dataset. For categorical variables, we present both the count and percentage of each category. For numerical variables, we summarize the data by reporting the mean and standard deviation, providing a clear sense of the central tendency and variability within the dataset.

**Table 1.** Demographics of pre-treatment features (PTF)

| **Group 1: Pre-treatment features** |  |  |  |
| --- | --- | --- | --- |
| Variable | All patients<br>(N=676) | Training set<br>(n=539) | Testing set<br>(n=137) |
| **age** | 60.49 (8.92) | 60.83 (9.10) | 59.15 (8.10) |
| **gender** |  |  |  |
| Male | 616 (91.1%) | 492 (91.3%) | 124 (90.5%) |
| Female | 60 (8.9%) | 47 (8.7%) | 13 (9.5%) |
| **ethnicity** |  |  |  |
| Not Hispanic or Latino | 612 (90.5%) | 483 (89.6%) | 129 (94.2%) |
| Hispanic or Latino | 49 (7.2%) | 44 (8.2%) | 5 (3.6%) |
| Unknown | 15 (2.2%) | 12 (2.2%) | 3 (2.2%) |
| **race** |  |  |  |
| White | 640 (94.7%) | 510 (94.6%) | 130 (94.9%) |
| Black or African American | 14 (2.1%) | 8 (1.5%) | 6 (4.4%) |
| Other Race | 17 (2.5%) | 16 (3.0%) | 1 (0.7%) |
| Asian | 4 (0.6%) | 4 (0.7%) | 0 (0.0%) |
| American Indian/Alaska Native | 1 (0.1%) | 1 (0.2%) | 0 (0.0%) |
| **smoking_status** |  | # Not Available |  |
| 4 | 290 (42.9%) | 243 (45.1%) | 47 (34.3%) |
| 5 | 344 (50.9%) | 267 (49.5%) | 77 (56.2%) |
| 1 | 26 (3.8%) | 20 (3.7%) | 6 (4.4%) |
| 2 | 5 (0.7%) | 3 (0.6%) | 2 (1.5%) |
| 7 | 6 (0.9%) | 3 (0.6%) | 3 (2.2%) |
| 9 | 2 (0.3%) | 1 (0.2%) | 1 (0.7%) |
| 10 | 3 (0.4%) | 2 (0.4%) | 1 (0.7%) |
| **tobacco_packs_per_day** | 0.65 (0.72) | 0.67 (0.73) | 0.58 (0.69) |
| **tobacco_used_years** | 13.49 (15.47) | 13.75 (15.68) | 12.49 (14.68) |
| **pack_years** | 14.99 (22.78) | 15.41 (23.46) | 13.32 (19.94) |
| **site_of_tumor** |  |  |  |
| BOT (base of tongue) | 337 (49.9%) | 272 (50.5%) | 65 (47.4%) |
| NOS (not ow specified) | 10 (1.5%) | 9 (1.7%) | 1 (0.7%) |
| Tonsil | 312 (46.2%) | 245 (45.5%) | 67 (48.9%) |
| Soft Palate | 8 (1.2%) | 8 (1.5%) | 0 (0.0%) |
| GPS | 6 (0.9%) | 2 (0.4%) | 4 (2.9%) |
| Pharyngeal Wall | 3 (0.4%) | 3 (0.6%) | 0 (0.0%) |
| **ajcc_version** |  |  |  |
| 7th Version | 48 (7.1%) | 40 (7.4%) | 8 (5.8%) |
| 8th Version | 628 (92.9%) | 499 (92.6%) | 129 (94.2%) |
| **p16_hpv_postive** |  |  |  |
| Yes | 596 (88.2%) | 478 (88.7%) | 118 (86.1%) |
| No | 56 (8.3%) | 41 (7.6%) | 15 (10.9%) |
| Unknown | 24 (3.6%) | 20 (3.7%) | 4 (2.9%) |
| **tumor_laterality** |  |  |  |
| Right | 345 (51.0%) | 270 (50.1%) | 75 (54.7%) |
| Left | 314 (46.4%) | 254 (47.1%) | 60 (43.8%) |
| Bilateral | 17 (2.5%) | 15 (2.8%) | 2 (1.5%) |
| **t_nominal** |  |  |  |
| t1 | 223 (32.8%) | 177 (32.7%) | 46 (33.6%) |
| t0 | 9 (1.3%) | 8 (1.5%) | 1 (0.7%) |
| t2 | 243 (35.9%) | 183 (34.0%) | 60 (43.8%) |
| t3 | 116 (17.0%) | 98 (18.0%) | 18 (13.1%) |
| t4 | 82 (12.1%) | 70 (13.0%) | 12 (8.8%) |
| tx | 3 (0.4%) | 3 (0.6%) | 0 (0.0%) |
| **n_nominal** |  |  |  |
| n2b | 35 (5.2%) | 28 (5.2%) | 7 (5.1%) |
| n1 | 446 (65.7%) | 355 (65.5%) | 91 (66.4%) |
| n2 | 94 (13.9%) | 78 (14.5%) | 16 (11.7%) |
| n0 | 54 (8.0%) | 42 (7.8%) | 12 (8.8%) |
| n2c | 23 (3.4%) | 17 (3.2%) | 6 (4.4%) |
| n3 | 18 (2.7%) | 14 (2.6%) | 4 (2.9%) |
| nx | 2 (0.3%) | 2 (0.4%) | 0 (0.0%) |
| n2a | 4 (0.6%) | 3 (0.6%) | 1 (0.7%) |
| **m** |  |  |  |
| m0 | 661 (97.8%) | 526 (97.6%) | 135 (98.5%) |
| mx | 10 (1.5%) | 8 (1.5%) | 2 (1.5%) |
| m1 | 5 (0.7%) | 5 (0.9%) | 0 (0.0%) |
| **status_at_enrollememt** |  |  |  |
| **Previously_Untreated** | 605 (89.5%) | 484 (89.8%) | 121 (88.3%) |
| **Previously_Treated** | 71 (10.5%) | 55 (10.2%) | 16 (11.7%) |

**Table 2.** Demographics of interventions/actions (ACTIONS)

| **Group 2: Actions** |  |  |  |
| --- | --- | --- | --- |
| Variable | All patients<br>(N=676) | Training set<br>(n=539) | Testing set<br>(n=137) |
| **definitive** |  |  |  |
| No | 609 (90.1%) | 490 (90.9%) | 119 (86.9%) |
| Yes | 67 (9.9%) | 49 (9.1%) | 18 (13.1%) |
| **induction** |  |  |  |
| No | 600 (88.8%) | 475 (88.1%) | 125 (91.2%) |
| Yes | 76 (11.2%) | 64 (11.9%) | 12 (8.8%) |
| **radio/chemo** |  |  |  |
| 2 Yes with concurrent<br>chemo? | 473 (70.0%) | 369 (68.5%) | 104 (75.9%) |
| 0 No | 29 (4.3%) | 26 (4.8%) | 3 (2.2%) |
| 1 Yes | 174 (25.7%) | 144 (26.7%) | 30 (21.9%) |

**Table 3.** Demographics of post-intervention outcomes (PIO)

| **Group 3: post-intervention outcomes** |  |  |  |
| --- | --- | --- | --- |
| Variable | All patients<br>(N=676) | Training set<br>(n=539) | Testing set<br>(n=137) |
| **ENE** |  |  |  |
| Positive | 50 (7.4%) | 38 (7.1%) | 12 (8.8%) |
| Negative | 13 (1.9%) | 7 (1.3%) | 6 (4.4%) |
| Unknown | 613 (90.6%) | 494 (91.6%) |  |
| **Margin** |  |  |  |
| Negative | 38 (5.6%) | 26 (4.8%) | 12 (8.8%) |
| Positive | 14 (2.1%) | 13 (2.4%) | 1 (0.7%) |
| <1mm | 3 (0.4%) | 3 (0.6%) | 0 (0.0%) |
| <2mm | 1 (0.1%) | 1 (0.2%) | 0 (0.0%) |
| <1m | 1 (0.1%) | 1 (0.2%) | 0 (0.0%) |
| <2m | 1 (0.1%) | 1 (0.2%) | 0 (0.0%) |
| 1mm | 1 (0.1%) | 1 (0.2%) | 0 (0.0%) |
| **rt_type** |  |  |  |
| IMPT | 165 (24.4%) | 137 (25.4%) | 28 (20.4%) |
| Vmat | 389 (57.5%) | 303 (56.2%) | 86 (62.8%) |
| IMRT | 105 (15.5%) | 83 (15.4%) | 22 (16.1%) |
| 3d Conformal | 2 (0.3%) | 2 (0.4%) | 0 (0.0%) |
| Other | 15 (2.2%) | 14 (2.6%) | 1 (0.7%) |
| **rt_dose** | 6823.85 (307.80) | 6829.26 (303.67) | 6802.55 (323.79) |
| **rt_fraction** | 32.56 (1.27) | 32.58 (1.30) | 32.50 (1.16) |

**Table 4.**
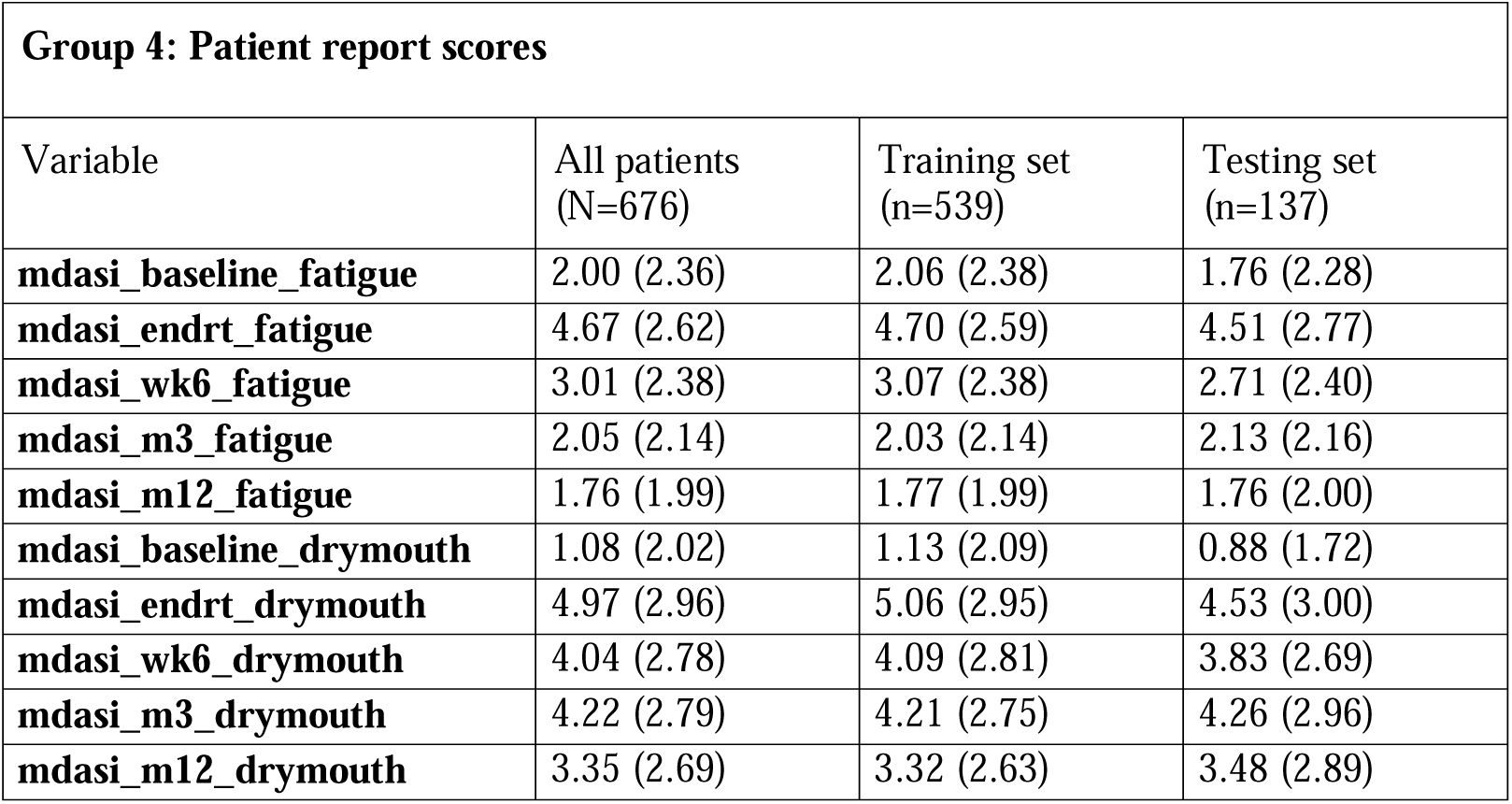
Demographics of patient report scores (PRS). The reported scores include the following metrics: pain, fatigue, nausea, sleep, distress, sob, memory, appetite, drowsy, drymouth, sad, vomit, numb, mucus, swallow, choke, voice, skin, constipation, taste, mucositis, teeth, activity, mood, work, relations, walking, enjoy. To save space, we only show the statistics of fatigue and drymouth. endrt means end of radiotherapy, wk6 means 6 weeks afterwards, and m3 means 3 months afterwards.

This summary offers a snapshot of the dataset’s structure.

## Temporal Modeling

### Case studies

To illustrate how our temporal process model can be utilized to formulate treatment plans and to clarify the rationale behind our feature categorization, we study two patients from the dataset as example cases. These cases demonstrate the diversity in treatment processes—one with a shorter treatment plan and the other with a more prolonged treatment history. By examining these cases, we can gain insight into how different treatment trajectories unfold in practice.

- Patient 1: This patient has a brief treatment history, involving one round of chemotherapy followed by radiotherapy. These treatments occurred close together, indicating a concurrent approach. Despite this aggressive intervention, the patient experienced a recurrence around 16 months later.

- Patient 2: This patient exhibits a more complex and prolonged treatment history, including multiple recurrences. The patient underwent various therapies such as radiation, surgery, and chemotherapy over an extended period. The disease shifted between periods of progression, stability, and partial remission, illustrating the need for a flexible, multi-modal treatment approach in managing recurring illness.

| Date | Category | Procedure |
| --- | --- | --- |
| 0 |  | Treatment Start |
| 28 | Action | Chemotherapy/Biotherapy/Investigational/SMI |
| 33 | Action | Radiation Therapy |
| 517 | Status | Relapse/Recurrence |

| Date | Category | Procedure |
| --- | --- | --- |
| 0 |  | Treatment Start |
| 53 | Action | Radiation Therapy |
| 168 | Status | No Evidence of Disease (NED) |
| 830 | Status | Relapse/Recurrence |
| 845 | Action | Surgery |
| 869 | Status | Progression |
| 888 | Action | Chemotherapy |
| 930 | Status | Stable Disease |
| 960 | Action | Surgery |
| 1027 | Action | Radiation Therapy |
| 1113 | Status | No Evidence of Disease (NED) |
| 2300 | Status | Progression |
| 2339 | Action | Surgery |
| 2489 | Action | Other Therapies |
| 2540 | Status | Partial Remission/Partial or Mixed Response |
| 2582 | Action | Other Therapies |
| 2624 | Status | Stable Disease |
| 2637 | Action | Surgery |
| 2666 | Action | Other Therapies |
| 2708 | Status | Stable Disease |

### Treatment as a temporal process

Based on the case studies, treatment can be broken down into the atomic segments of

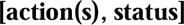

Take patient 1 as example, this patient has

- Action 1 as Chemotherapy and action 2 as Radiation, and due to their proximity between the dates, they are considered as an action of radiotherapy (w/ concurrent chemotherapy)
- Status 1 is relapse at day 517 but no follow-up action is taken.

The [actions, status] segment becomes a cycle of treatment.

And multiple cycles of this pattern can occur if relapse or recurrence happens. For instance, a treatment course might look like

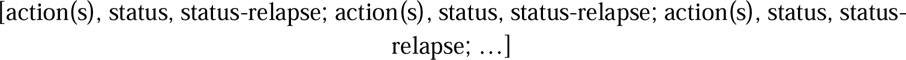

with each cycle separated by a semicolon. The actions in each cycle can vary, depending on the patient’s response and disease progression.

The patient 2 is an example of receiving multiple cycles.

### Simplified modeling

This multi-cycle approach, however, poses challenges for modeling, as the treatment plan can vary in length and, in theory, can extend indefinitely. To strike a balance between modeling accuracy and practical application, we simplify this multi-cycle treatment into a single-cycle model. This is illustrated by the following example:

In addition, we only consider the outcome up to year 3, as a variable of relapse_yr3 as shown in Table 5.

**Table 5.** Demographics of treatment plan outcomes (TPO)

| **Group 5: Treatment plan outcomes** |  |  |  |
| --- | --- | --- | --- |
| Variable | All patients<br>(N=676) | Training set<br>(n=539) | Testing set<br>(n=137) |
| **relapse_yr3** |  |  |  |
| False | 153 (22.6%) | 123 (22.8%) | 30 (21.9%) |
| True | 35 (5.2%) | 29 (5.4%) | 6 (4.4%) |

[action_1_, status_1_, status-relapse; action_2_, status_2_, status-relapse; action_3_, status_final_] is simplified to [action_1_, status_yr3_]. See the resulted patient 1 and patient 2 below as examples.

In this simplified approach, we only model the first cycle of treatment and retain the year 3 status as the outcome. The rationale for this simplification is threefold: (i) it ensures a fixed length of variables, making the model more manageable; (ii) the final status serves as a measure of the overall effectiveness of the entire treatment plan; and (iii) the intermediate actions and statuses can be viewed as outcomes of the initial action(s), functioning as hidden or latent variables from a probabilistic point of view. Revisiting our example patients, their treatment plans are now modeled as follows:

- Patient 1: Remain unchanged.

- Patient 2: Now modeled as

| Date | Category | Procedure |
| --- | --- | --- |
| 0 |  | Treatment Start |
| 28 | Action (cycle 1) | Chemotherapy/Biotherapy/Investigational/SMI |
| 33 | Action (cycle 1) | Radiation Therapy |
| 517 | Status (final) | Relapse/Recurrence |

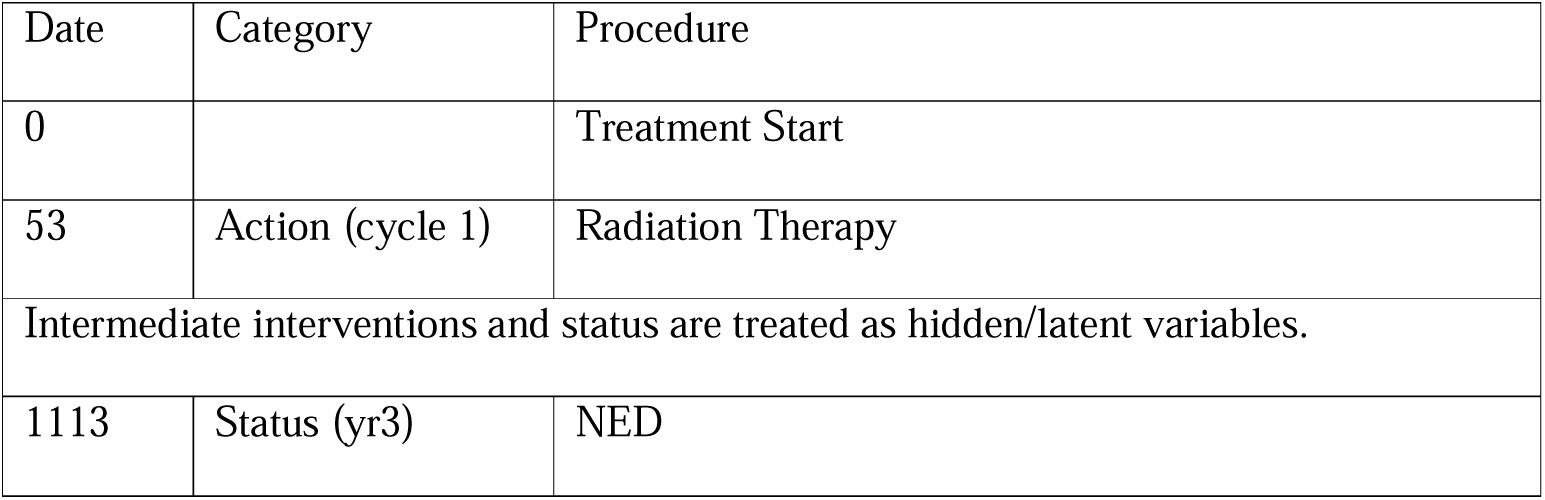

### First-cycle interventions

While multi-cycle treatments have been simplified to single-cycle models—intermediate interventions treated as latent variables—the first cycle itself can still involve various interventions. Patient #1 above is an example. More critically, the number and order of these interventions are not fixed. Modeling a variable number of interventions with dynamic ordering can be complex. However, our analysis of the dataset reveals that treatment plans generally follow a certain pattern devised by doctors. By applying specific data-cleaning and preprocessing steps, combined with the first-cycle simplification, we identified a consistent pattern in most patients’ treatment sequences, as shown below:

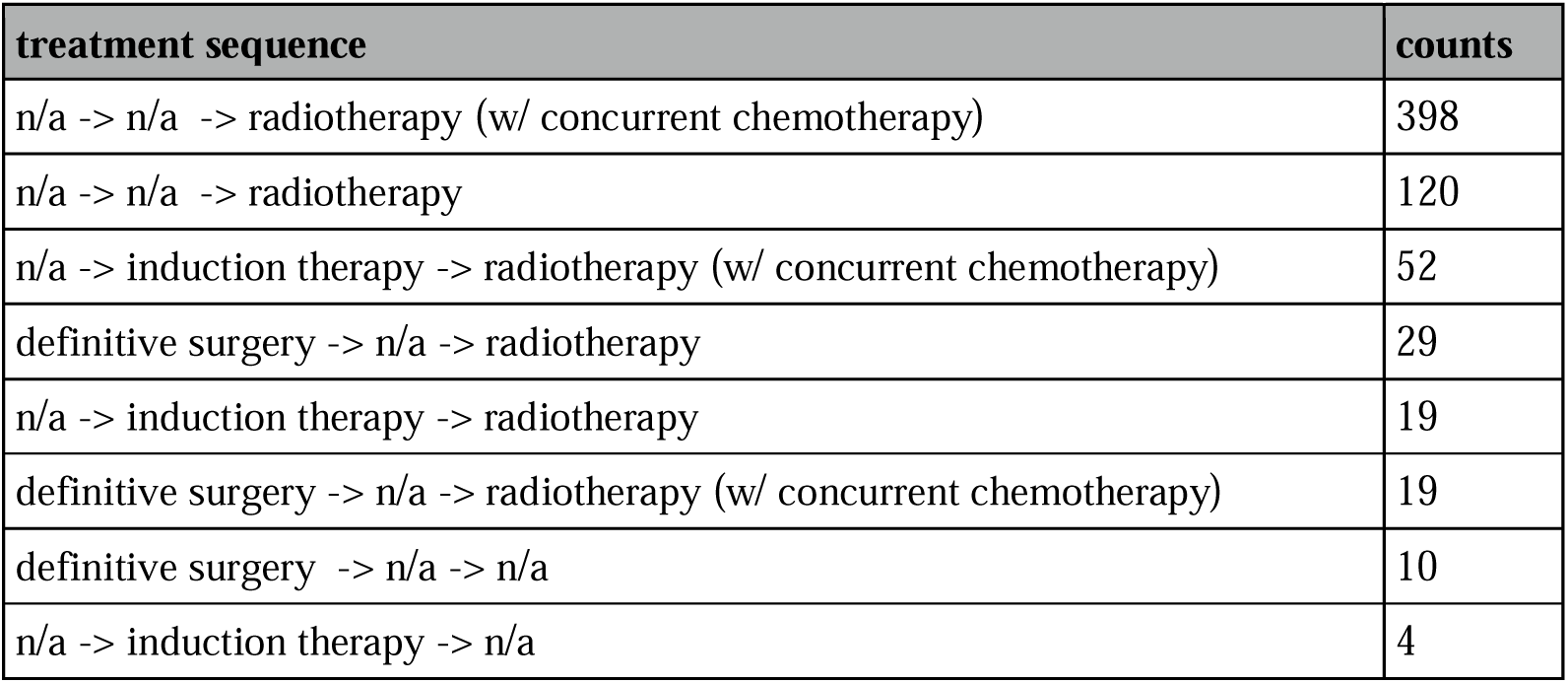

The patterns mined from the data suggest that the majority of patients can be effectively modeled using the following sequence of variables, with only a few exceptions:

- Action 1: Definitive Surgery (binary variable – no/yes)
- Action 2: Induction Therapy (binary variable – no/yes)
- Action 3: Radiotherapy (three classes – no/yes/yes-with-concurrent-chemotherapy)

Based on these action variables, the treatment sequence can now be formulated into a structured diagram, simplifying the modeling process by capturing the essence of the first-cycle interventions.

Therefore, with our single-cycle formulation and pattern mining, we have established a fixed pattern of treatment sequences that can be effectively captured and predicted. This structured approach allows us to model these treatment plans using conventional machine learning techniques, bypassing the complexities associated with variable-length and dynamic-order interventions.

## Simulator Details

### Overview

Building on our sequential modeling approach and the duo-component design, our simulator consists of: (i) a patient generator, which functions as a generative component to produce patient features before any treatment, and (ii) a development predictor, a predictive component that estimates the step-by-step outcomes of interventions for a given patient.

### Patient Generator

We implemented a customized Variational Autoencoder (VAE) tailored to handle heterogeneous patient data, encompassing both categorical and numerical features. The VAE is designed to generate a realistic representation of patient characteristics prior to any treatment, drawing samples that reflect the complex distributions found in the population. By capturing this pre-treatment patient status, the VAE ensures a robust starting point for simulating various clinical scenarios.

### Development Predictor

To forecast the progression of disease and the effects of subsequent treatments, we utilized XGBoost as the core of our predictive model. Unlike the patient generator, the development predictor must account for the sequential nature of patient data, where future outcomes depend on prior states and interventions. XGBoost, known for its efficiency and high predictive accuracy in complex datasets, is well-suited for this task. It systematically models the step-by-step evolution of a patient’s condition, using a gradient-boosting approach to handle the intricate relationships between clinical actions and patient responses. This enables the predictor to capture nuanced interactions and predict future developments, even with heterogeneous features and a varying number of classes for categorical interventions.

In our design, the generative VAE creates the initial patient features, assuming they are independently drawn from underlying distributions. However, as treatments are administered and the disease progresses, these developments follow a sequential pattern. This sequential dependency makes XGBoost an ideal choice for the development predictor, as it can effectively model the trajectory of a patient’s condition over time by learning from previous states and interventions. Figure 2 as well as Figure 3 provide a schematic of our simulator, illustrating the integration of our sequential modeling approach to simulate patient outcomes dynamically.

**Figure 2.**
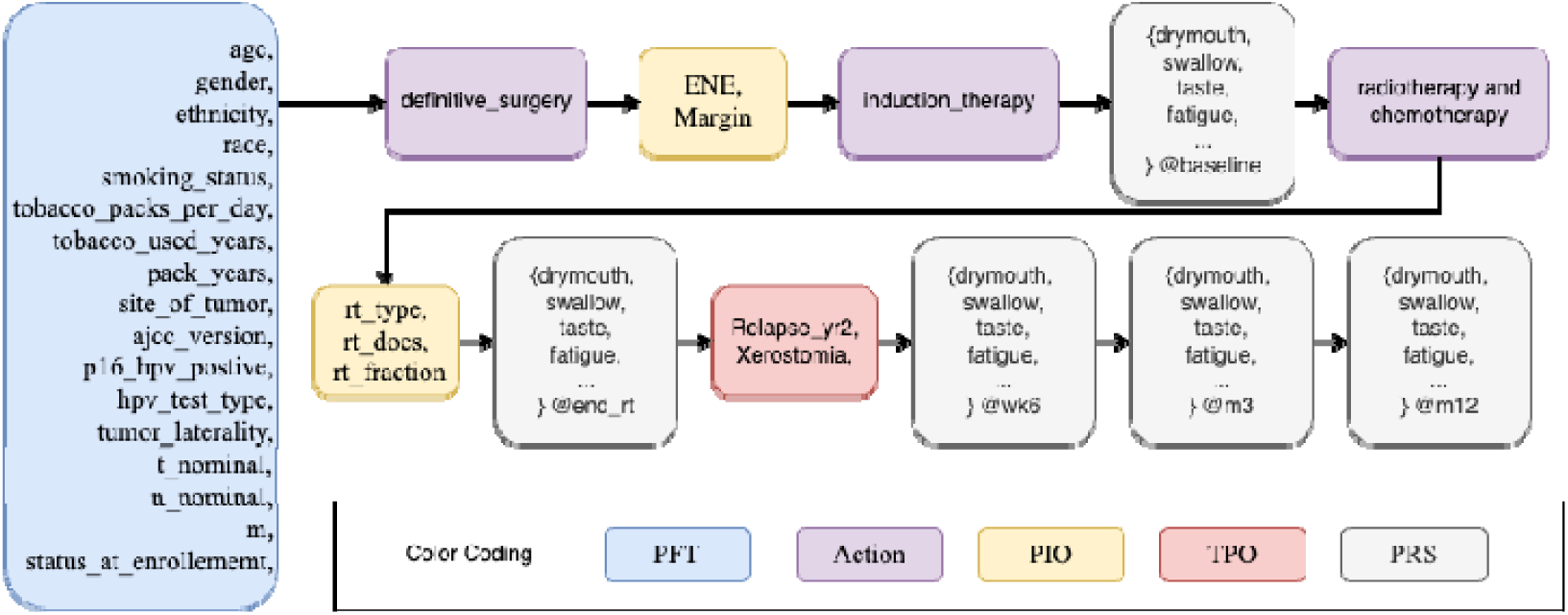
Temporal modeling diagram of treatment process.

**Figure 3.**
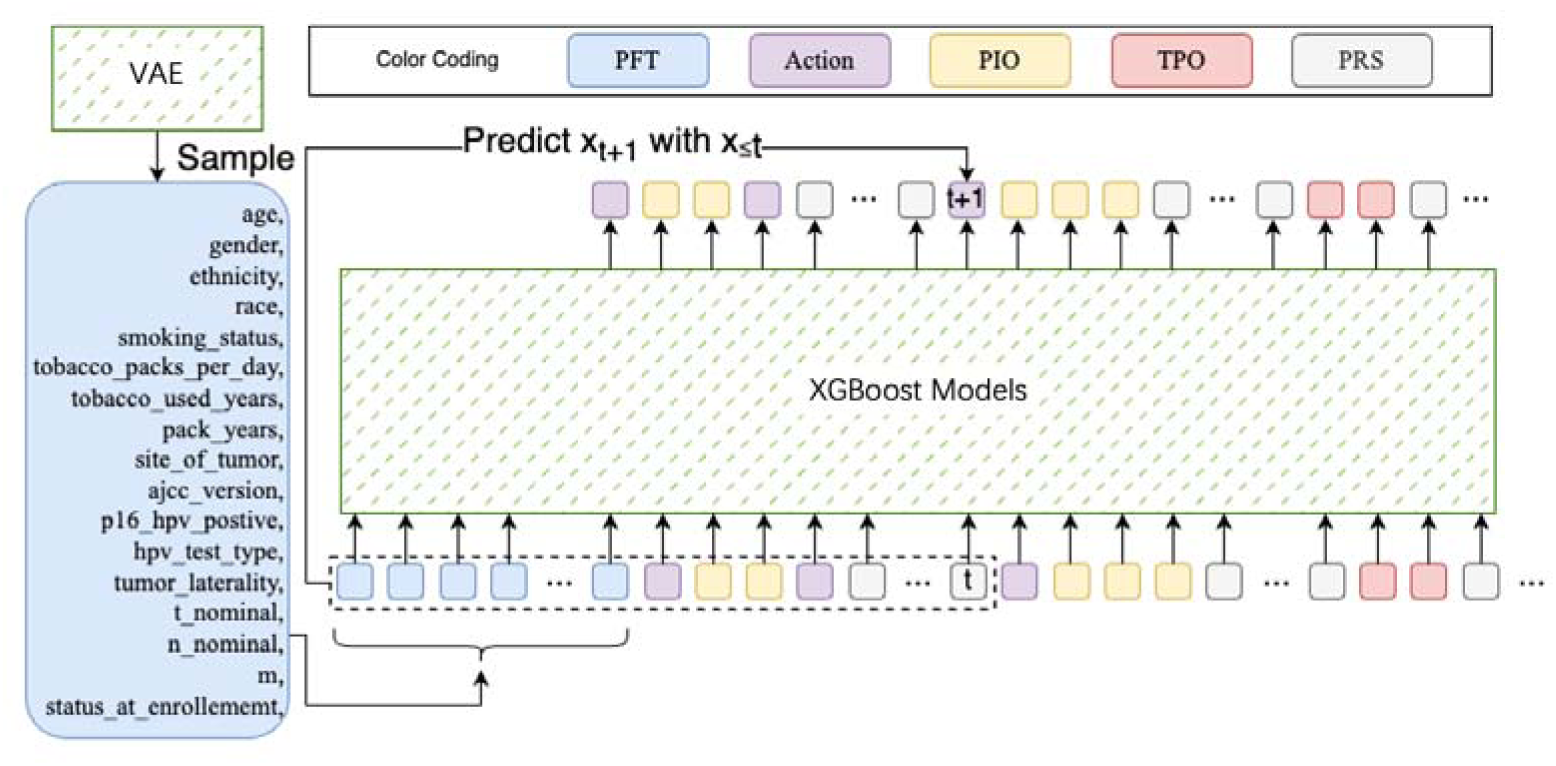
Simulator details.

### VAE Preliminary

Variational Autoencoders (VAEs) offer a probabilistic approach to generating new patient data by learning the underlying patterns in complex datasets. They work by introducing hidden “latent variables” that capture the essential characteristics of patient features, as shown in Figure 4.

**Figure 4.**
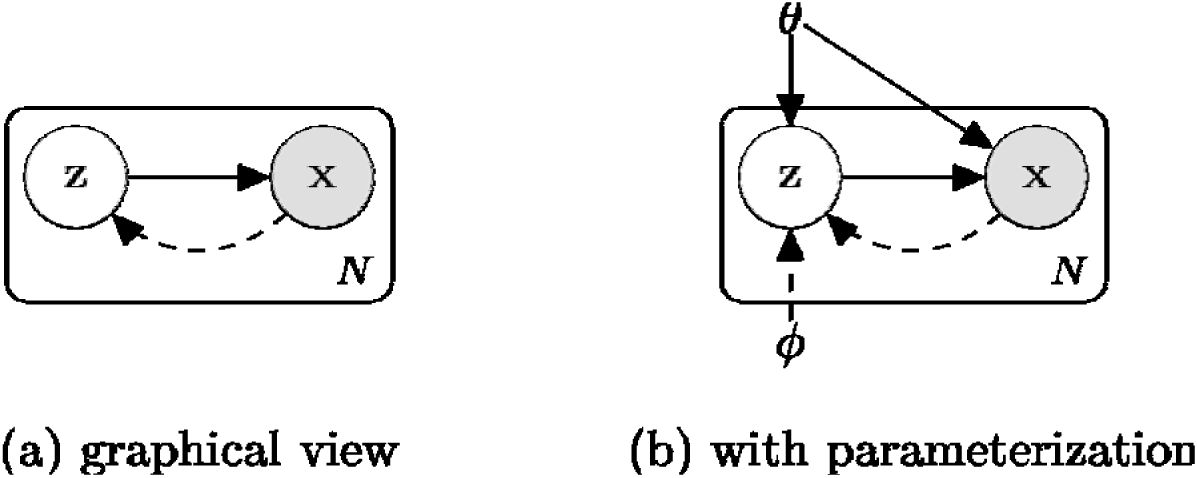
A probabilistic view of VAE

Imagine a set of patients, each with various characteristics like age, symptoms, and test results. Directly sampling from this complex distribution is challenging. VAEs address this by assuming that the patient data can be generated through a simpler process involving latent variables. These variables represent the underlying factors that influence the observed patient data.

The VAE framework has two key components:

- **Encoder**: This compresses the patient data into the latent space, capturing the essential patterns. It provides a mapping from the observed patient features to the latent variables.
- **Decoder**: This takes the latent variables and reconstructs them into the original patient space, generating new patient data that resembles the original dataset.

To train the VAE, we use an optimization objective known as the Evidence Lower Bound (ELBO). This objective consists of two parts:

- Reconstruction Term: Measures how well the model can recover the original patient data from the latent variables.
- Regularization Term: Ensures that the latent variables follow a smooth and tractable distribution, like a Gaussian. This makes it easier to generate new samples from the latent space.

The full optimization objective can be written as:

In simpler terms, this equation helps the model find a balance between accurately reconstructing the patient data and maintaining a well-behaved latent space. By maximizing this objective, the VAE learns to approximate the true data distribution in a way that’s easy to sample from. This is crucial for our simulator, as it allows us to generate realistic patient profiles before any treatment interventions.

By training the VAE in this way, we ensure that the latent variables capture the underlying diversity of patient data, making it possible to simulate various clinical scenarios realistically. This process of encoding and decoding forms the foundation of our simulator, allowing us to explore how different interventions might influence disease progression.

### XGBoost

XGBoost is a powerful and efficient algorithm widely used for predictive modeling tasks, especially when dealing with large and complex datasets like those in healthcare. In simple terms, it is an advanced form of decision tree-based learning that iteratively builds a series of decision trees to improve the model’s accuracy.

XGBoost works by constructing an ensemble of decision trees in a sequential manner. Each tree is built to correct the errors of the previous ones. This way, the model learns complex relationships in the data by focusing on the mistakes made by the earlier trees. The “boosting” part of XGBoost refers to this process of improving the model by combining multiple weak learners (decision trees) into a strong one, as Figure 5 shown.

**Figure 5.**
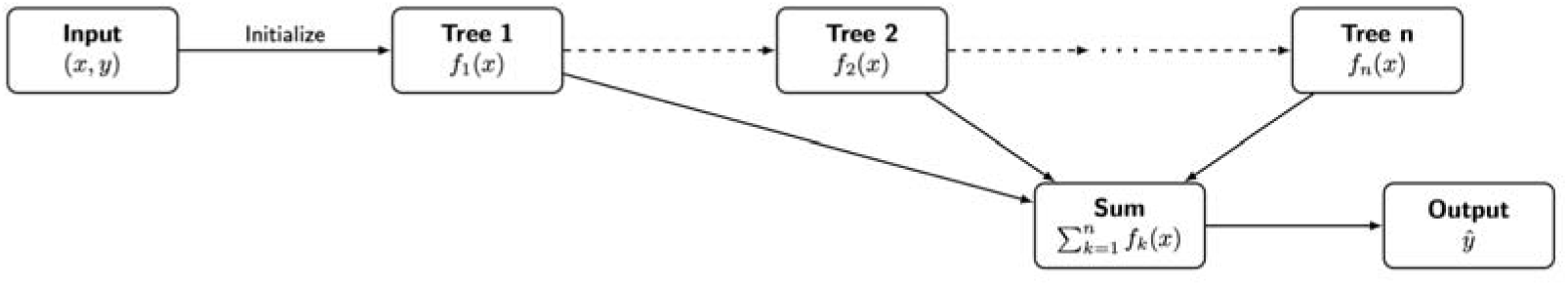
Overview of XGBoost.

The objective of XGBoost can be broken down into two main parts:

- Prediction Accuracy: This measures how well the model’s predictions match the actual outcomes. The goal is to minimize the difference between the predicted values and the true values.
- Regularization: This term prevents the model from becoming too complex, which can lead to overfitting. Regularization helps to ensure that the model generalizes well to new, unseen data by controlling the complexity of the individual trees.

The overall optimization objective of XGBoost can be expressed as:

- The first part represents the loss function, which measures how accurate the predictions \hat{y}_i are compared to the actual values y_i. This encourages the model to make predictions that closely match the real outcomes.
- The second part represents the regularization term, which controls the complexity of each decision tree f_k. This term helps to prevent overfitting by penalizing overly complex models.

XGBoost uses a gradient boosting approach to optimize this objective. It builds each new tree to minimize the errors of the current ensemble of trees. By focusing on the residuals—the differences between the actual and predicted values—XGBoost iteratively improves the model, capturing complex patterns in the data.

In our simulator, XGBoost serves as the predictive component that models the progression of disease and the effects of different treatments. Its ability to handle heterogeneous features and missing data, along with its robustness in capturing complex relationships, makes it well-suited for forecasting patient outcomes. By leveraging the sequential learning process of XGBoost, the simulator can make accurate step-by-step predictions about how different interventions might influence a patient’s condition over time.

## Simulator Results

### Generative component (VAE)

Recall that we utilize a Variational Autoencoder (VAE) as the generative model to simulate patients and their pretreatment features, ensuring that the generated data aligns with the real data distribution.

To evaluate the performance of the VAE, we generate 500 synthetic patients and use two complementary approaches for assessment:

- Visual comparison: We compare, in Figure 7, the distribution of each individual feature between the synthetic and real data. By plotting these feature-wise distributions side by side, we can visually inspect how well the VAE reproduces the patterns and variability present in the original dataset.
- Statistical distance: To quantitatively measure the similarity between the generated and real data, we compute the Wasserstein distance for each feature independently, shown in Figure 6. The feature-wise distances are averaged to provide an overall metric of alignment between the synthetic and real patient data, offering a robust statistical measure of the VAE’s generative accuracy.

**Figure 6.**
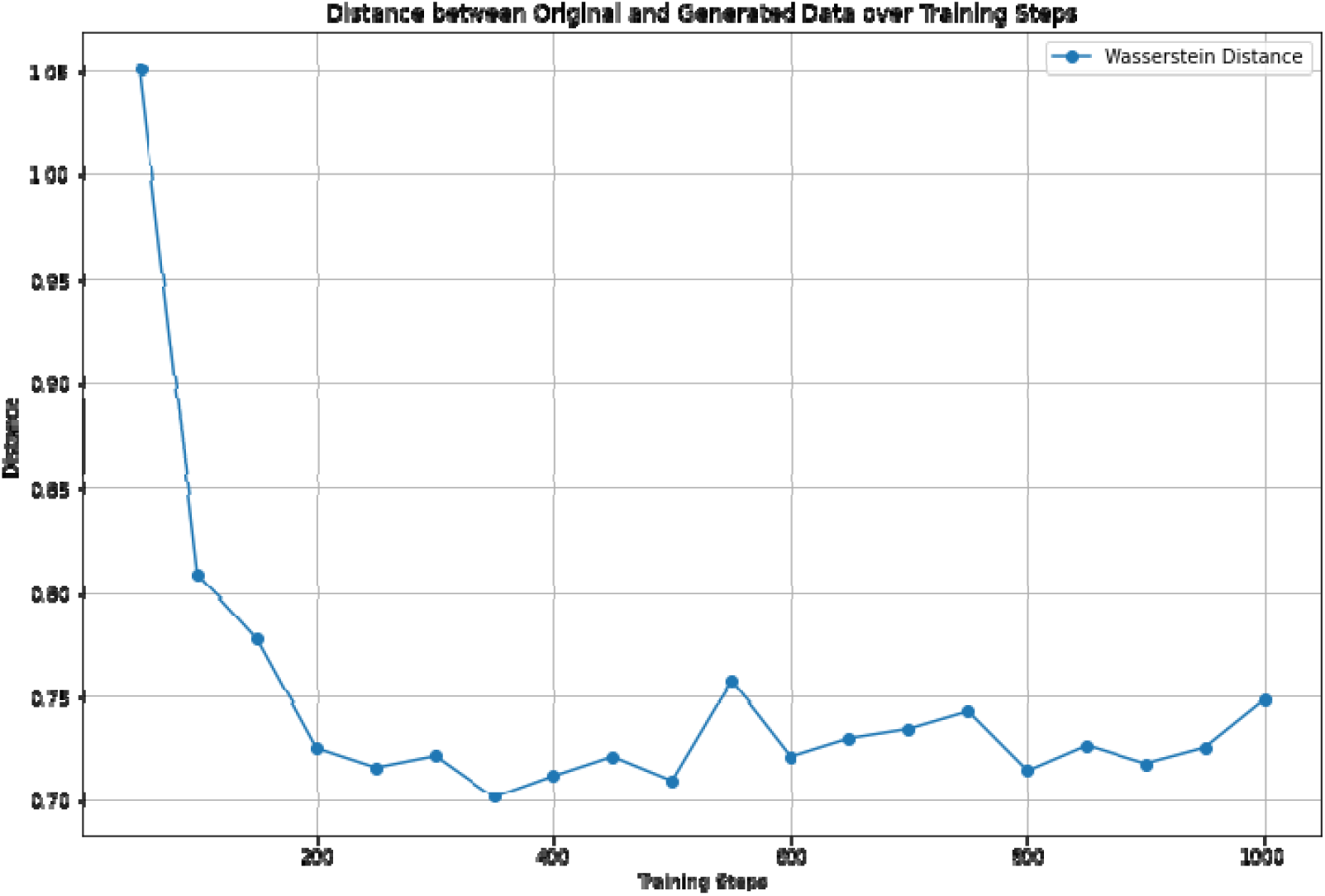
Wasserstein distance between synthetic patients and real patients during training.

**Figure 7.**
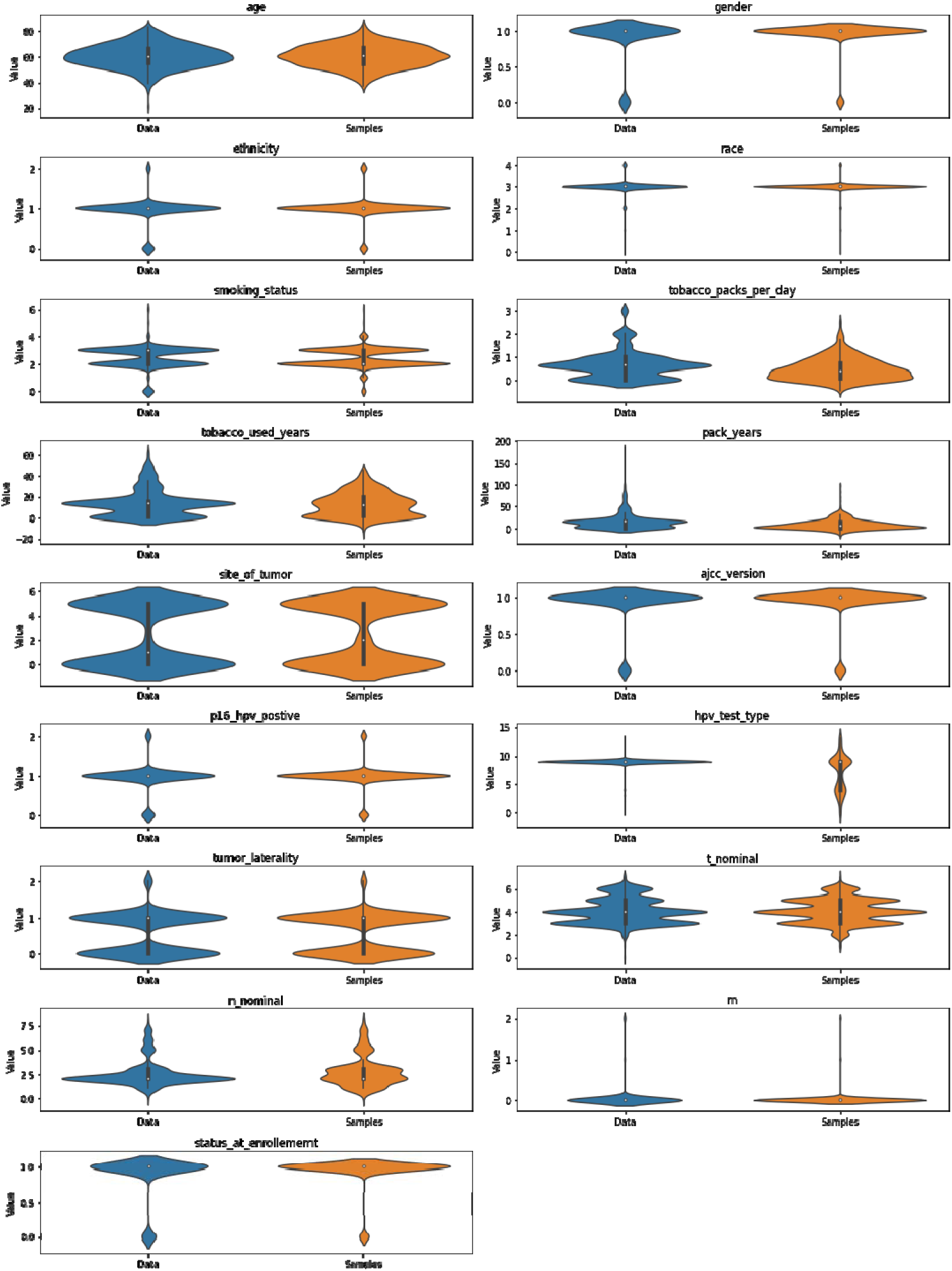
Feature-wise violin plot comparing the distribution of synthetic patients and real patients.

### Predictive component (XGBoost)

To evaluate the predictive performance of our model, we trained an XGBoost model and compared its results against several baselines, including Median/Mean values, a Random model, and a Multi-Layer Perceptron (MLP) model. The comparison was conducted across a range of features using two key metrics: the macro F1 score for classification tasks and Mean Squared Error (MSE) for regression tasks.

### Baselines

- Median/Mean Baseline: For classification tasks, this baseline involves predicting the median value for categorical features, while for numerical features, it simply predicts the mean value.
- Random Baseline: This baseline generates predictions randomly on the test set. For categorical features, the random predictions are proportional to the label distributions observed in the training set, providing a simple but probabilistically informed benchmark.
- MLP: Serving as a learning baseline.

### Overall Performance

- Macro F1 Score (↑ the higher the better): Given the highly imbalanced nature of the dataset, the macro F1 score was used to better capture the performance across all classes. The XGBoost model consistently outperformed these baselines across various classification tasks, achieving higher macro F1 scores. This indicates that XGBoost made more accurate and balanced predictions across all classes, effectively handling the imbalance in the data.
- Mean Squared Error (↓ the lower the better): For regression tasks, the XGBoost model achieved significantly lower MSE values than both the Median/Mean and Random baselines, indicating more precise predictions for continuous outcomes.

To save some space, we only take four reporting: fatigue, drymouth, swallow, and taste for experimenting. And only show the visualization (Figure 8, Figure 9, Figure 10 and Figure 11) for swallow (rest could be found in appendix).

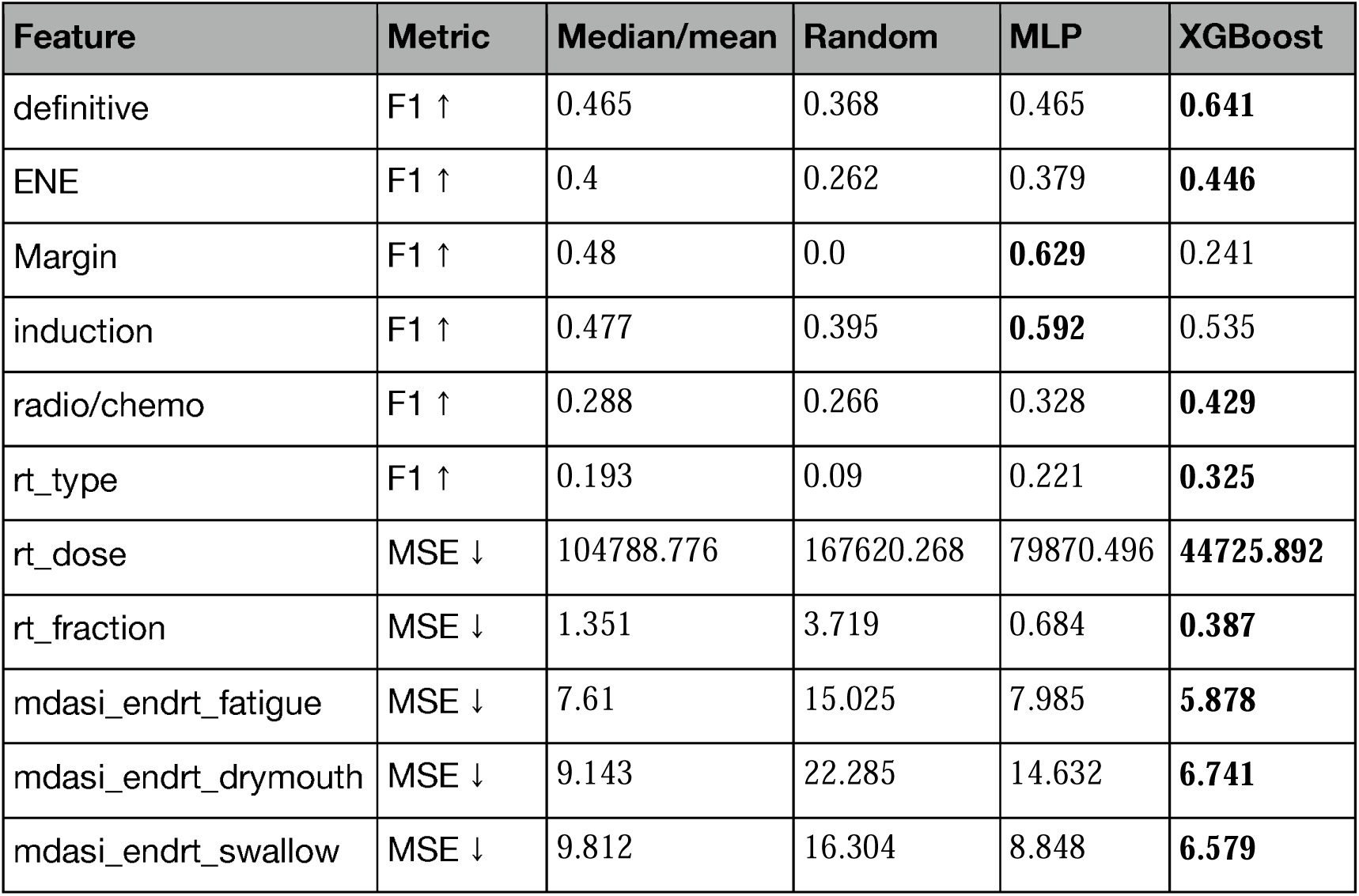

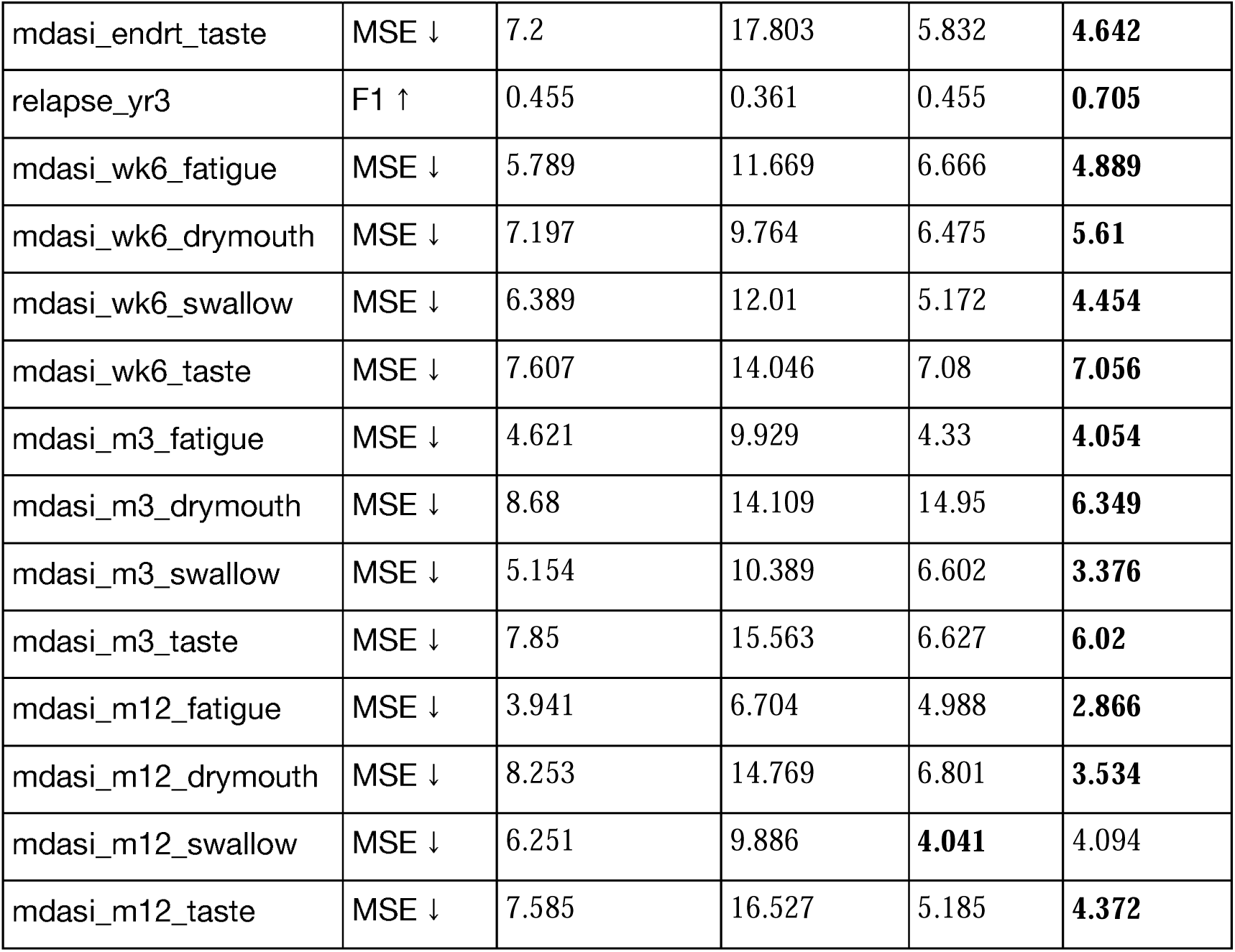

**Figure 8.**
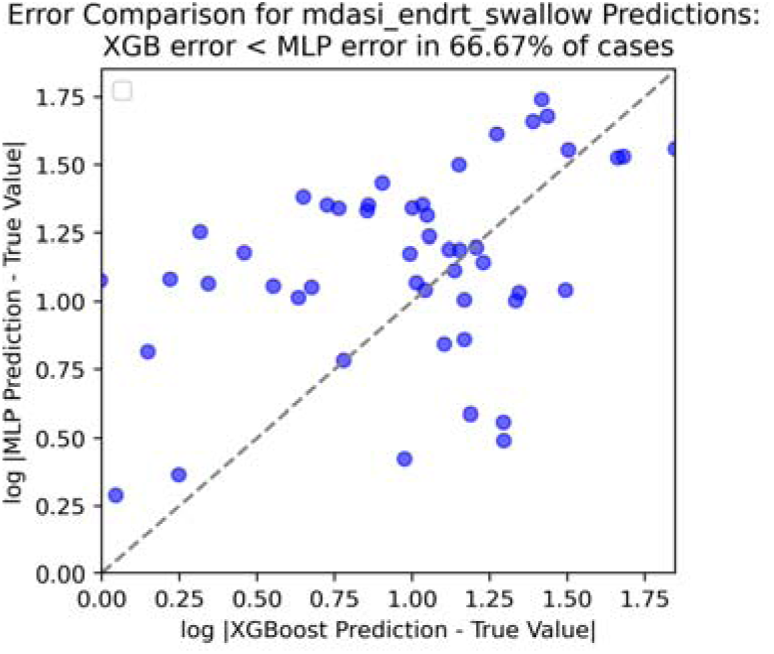
Error comparison for swallow score at end of RT.

**Figure 9.**
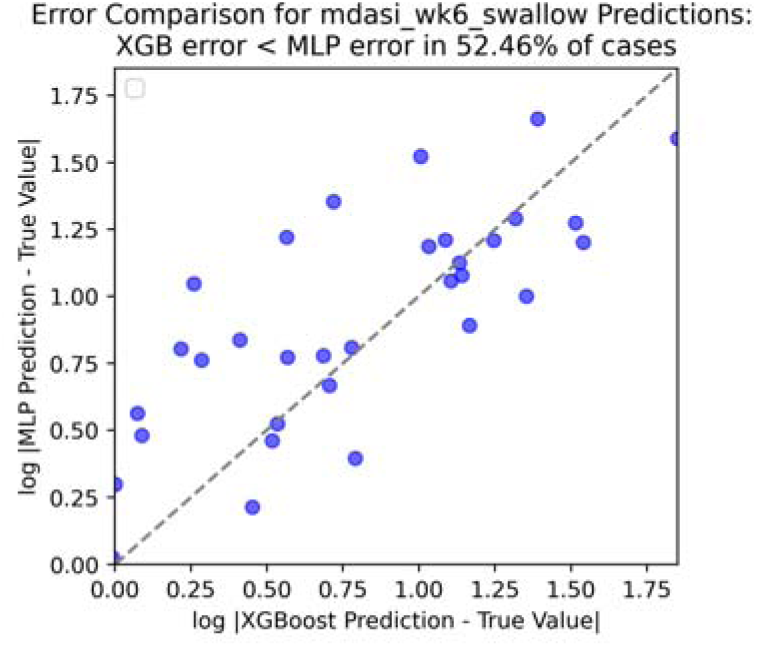
Error comparison for swallow score at end of week 6.

**Figure 10.**
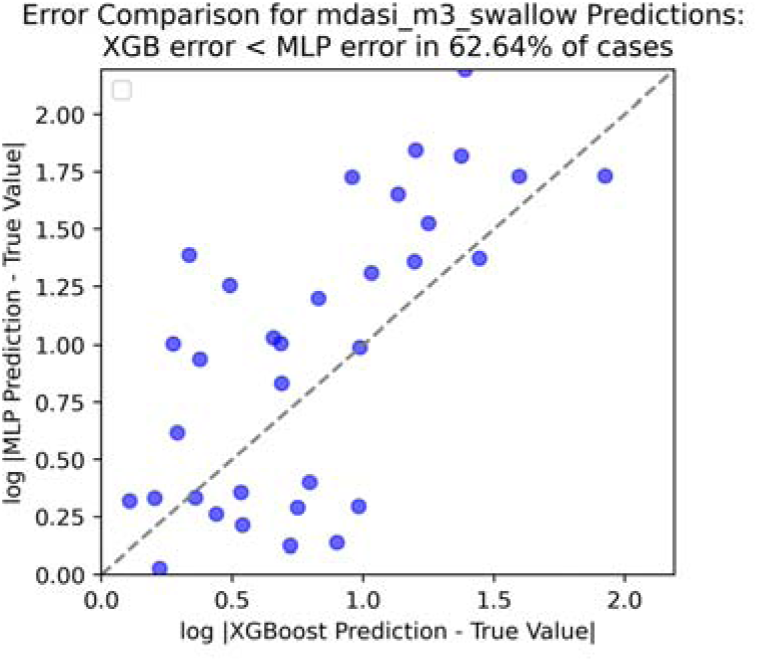
Error comparison for swallow score at end of month 3.

**Figure 11.**
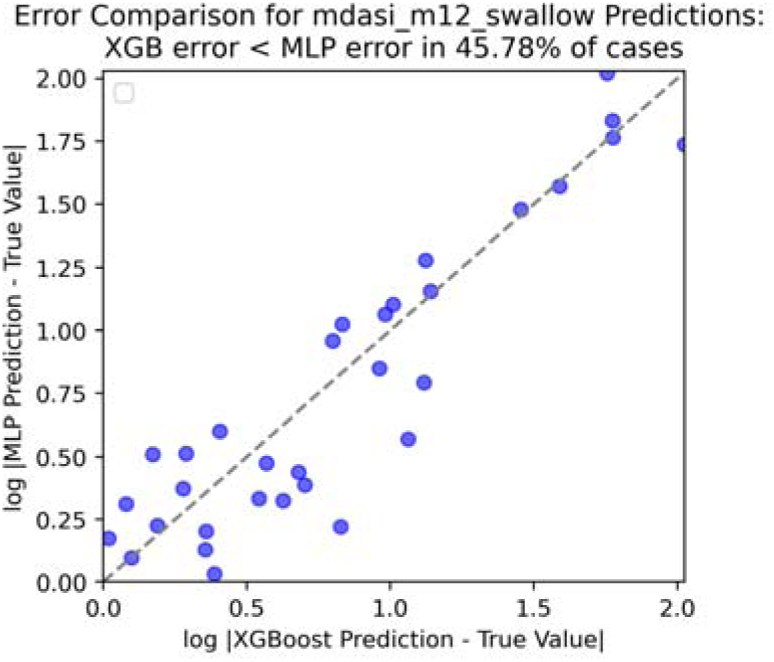
Error comparison for swallow score at end of month 12.

## Implementation Details

In this section, we explain the implementation details of our simulator, including data preprocessing, training details, handling heterogeneous inputs/outputs, etc.

### Data preprocessing

Following standard machine learning techniques, we apply data normalization, missing value imputation, train/test split to our dataset.

- Data normalization: We normalize all features to normal distribution with zero mean, unit standard deviation. In this case, the output is also expected to be normalized, we also demoralize the outputs with the recorded dataset mean and standard deviation.
- Missing value imputation: We handle missing values differently for the VAE model and the Transformer model. For VAE and MLP models, we use mean imputation for numerical features, and median imputation for categorical features. For the XGBoost models, we do not impute any values, as XGBoost is designed to be compatible with missing values.
- Train/test split: we randomly split the dataset into 539 train patients and 137 test patients.

### VAE training

The training of the Variational Autoencoder (VAE) consists of two key tasks: reconstructing the input data and regularizing the latent space. The model is designed to learn both numerical and categorical features from patient data. For numerical features, reconstruction is achieved using mean squared error (MSE), while categorical features are handled with cross-entropy loss. The VAE also employs Kullback-Leibler (KL) divergence to regularize the latent space, ensuring that it follows a standard normal distribution. This allows the VAE to generate meaningful synthetic data that aligns with the real data distribution.

The VAE was trained using the Adam optimizer with a learning rate of 0.001 and a batch size of 64, over 10,000 epochs. The encoder consists of two fully connected layers with 64 hidden units and ReLU activation, followed by two heads to calculate the mean and variance of the latent space, which has 16 dimensions. The decoder mirrors the encoder’s architecture, reconstructing the numerical and categorical data through separate output layers to match the input structure.

### XGBoost Training

For the XGBoost models, both classification and regression tasks are handled differently based on the nature of the target feature (categorical or numerical).

- Classification: We use the XGBClassifier with the objective set to ‘multi:softmax’ for multi-class classification and ‘binary:logistic’ for binary classification. The evaluation metric is set to ‘mlogloss’. Sample weights are computed using a balanced class weighting strategy to handle class imbalance. StratifiedKFold cross-validation is employed with 3 splits to ensure robust evaluation.
- Regression: For numerical target features, the XGBRegressor is used with the ‘neg_mean_squared_error’ as the scoring metric.

### MLP Training

The MLP model is used for both classification and regression tasks. For categorical targets, the MLPClassifier is trained using a grid search with 3-fold cross-validation, and the evaluation metric is set to ‘f1_macro’. For numerical targets, the MLPRegressor is used, with ‘neg_mean_squared_error’ as the scoring metric.

### Grid Search and Hyperparameters

Grid search is used to optimize the hyperparameters of both XGBoost and MLP models. The grid search was performed over the following ranges using a 3-fold cross-validation:

- For XGBoost:
  - learning_rate: [0.01, 0.1, 0.3]
  - max_depth: [4, 6, 8]
  - min_child_weight: [1, 3, 5]
  - n_estimators: [100, 200, 300]
  - colsample_bytree: [0.3, 0.5]
- For MLP:
  - hidden_layer_sizes: [(50, 50), (100, 100), (100, 50)]
  - activation: [‘relu’]
  - solver: [‘adam’, ‘sgd’]
  - learning_rate_init: [0.001, 0.01]
  - max_iter: [200, 400]

## Gym Wrapper

The OpenAI Gym provides a flexible toolkit for developing and comparing reinforcement learning (RL) algorithms by creating environments with defined actions, states, and rewards. In our implementation, we encapsulate our trained Variational Autoencoder (VAE) and XGBoost models within a custom Gym environment (MdasiGym). This environment simulates a clinical decision-making process over three key treatment steps. By wrapping the models in this Gym environment, we enable a plug-and-play interface where RL algorithms can interact with the models seamlessly. Researchers can train their RL models to make treatment decisions efficiently without needing to directly manage the underlying predictive models, making this setup both practical and adaptable for various clinical applications.

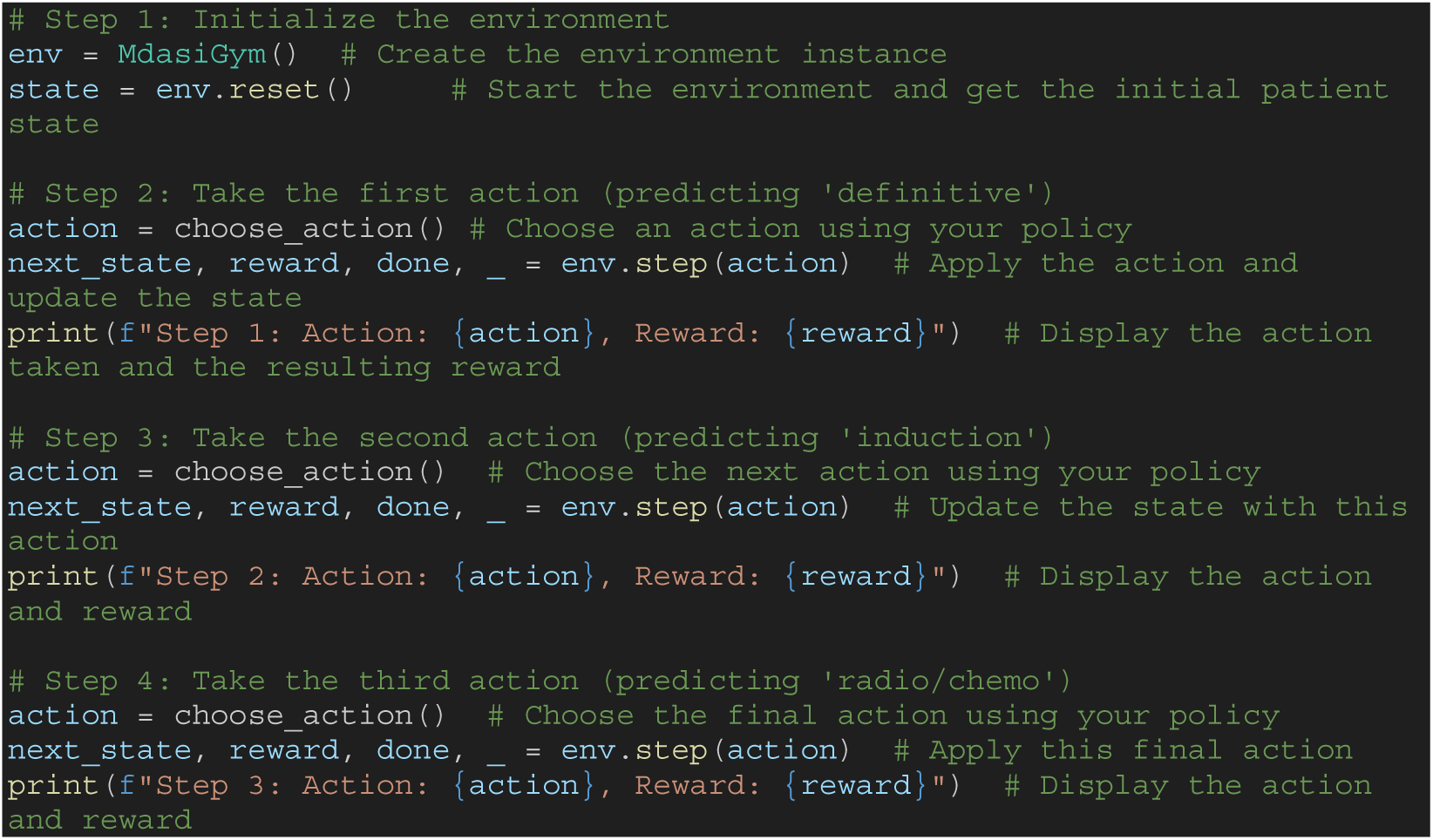

To demonstrate how our simulator simulates disease progression, we test two extreme of treatment plans and the results are presented in Figure 12:

- Plan 1: No definitive surgery, no induction, radiotherapy without chemotherapy
- Plan 2: Definitive surgery, induction, radiotherapy with chemotherapy

**Figure 12.**
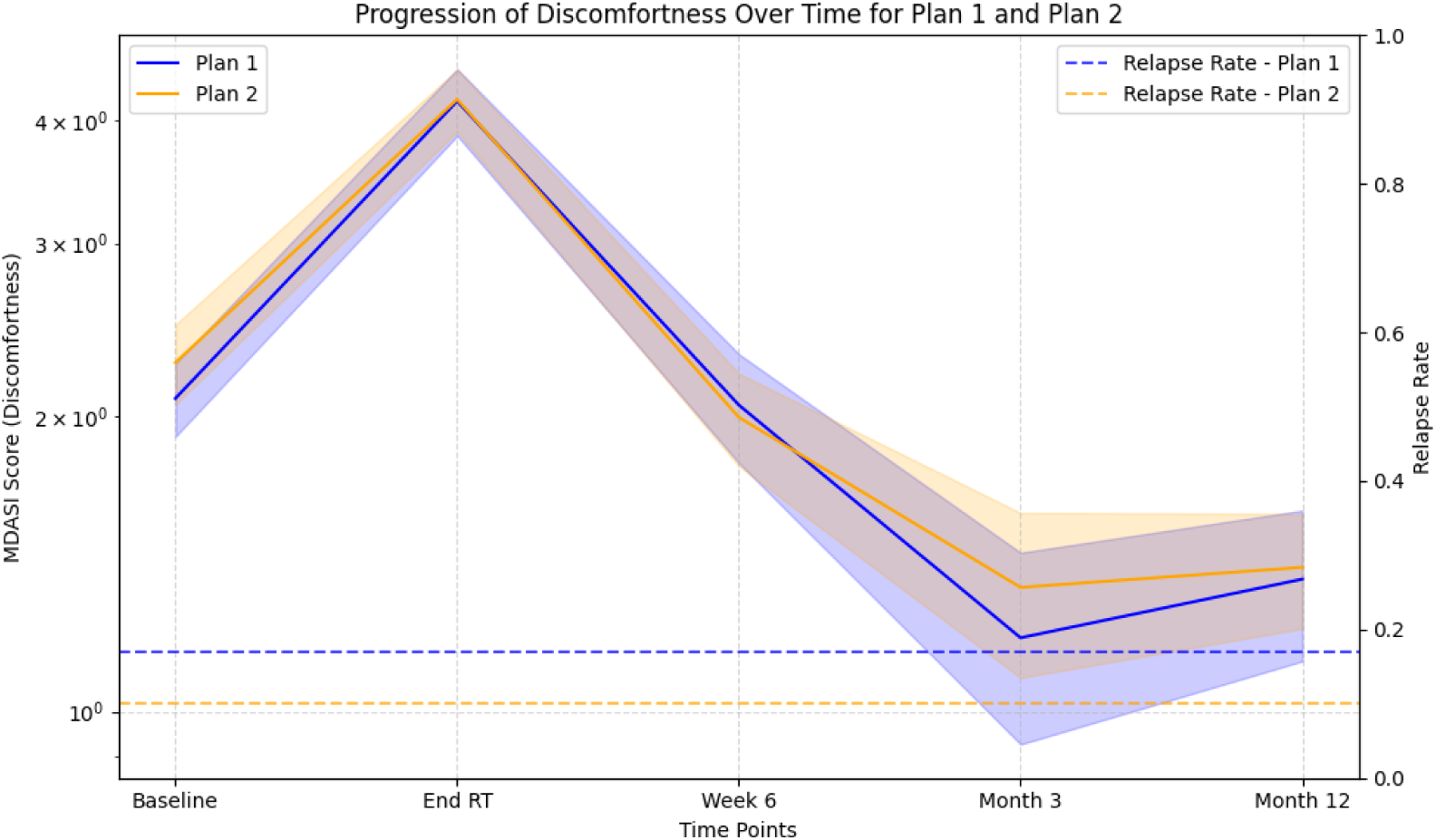
Disease progression prediction for two extreme treatment plans (Plan 1 is conservative; Plan 2 is aggressive): plan 2 leads to a lower relapse rate but slightly greater discomfort.

### Contribution Statement

- Conceptualization - formulation research ideas and aims: Z.Y., X.Z., G.C., C.D.F., G.E.M., A.W., C.F., Y.W., S.K.A.
- Methodology - design and development of methodology: Z.Y., X.Z., G.C.
- Software - implementation of program: Z.Y.
- Data Curation: S.K.A., A.S.R.M., M.N., C.D.F.
- Writing: Z.Y., X.Z., G.C., A.W., Y.W., H.J.H.
- Supervision: X.Z., G.C., C.D.F., G.E.M.
- Funding acquisition: X.Z., G.C., C.D.F., G.E.M.

### Ethics and Consent to Participate declarations

This retrospective study was exempt under MD Anderson IRB protocol RCR-003-0800. In compliance with the Health Insurance Portability and Accountability Act (HIPAA), informed consent was waived and approved by the IRB as all analyses were performed over retrospective anonymized data.

## Data Availability

The clinical dataset comprises retrospective patient records from the MD Anderson Cancer Center and cannot be publicly shared due to institutional restrictions. Aggregated statistics and variable summaries are included in the manuscript (Tables 1 to 5). De-identified synthetic patient data generated by the simulator and analysis code will be made available from the corresponding authors upon reasonable request and, where applicable, subject to MDACC data use agreements.

## Appendix

## Training dynamics of VAE

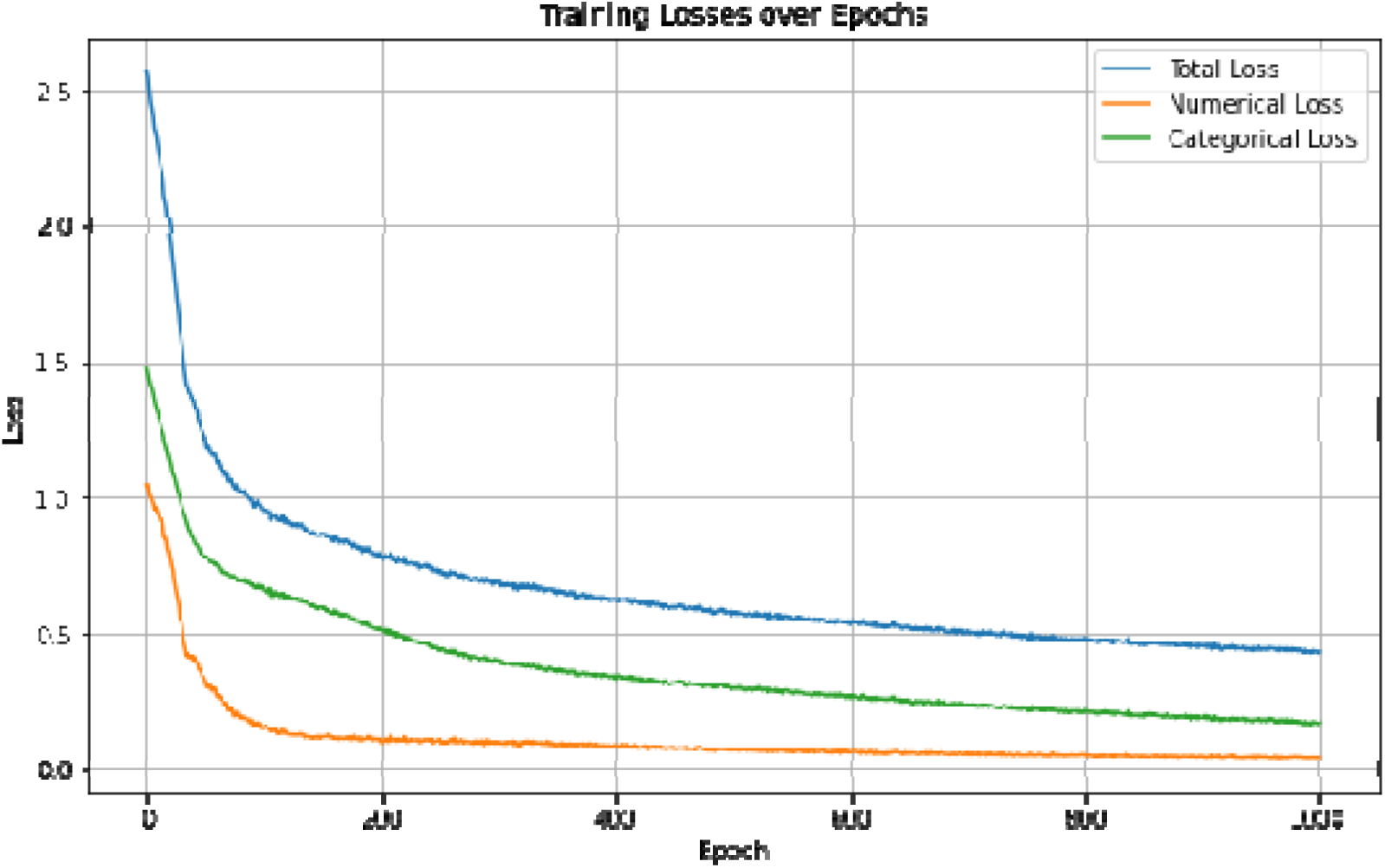

The loss curves during training demonstrated a clear downward trend in the total loss, as well as the individual numerical, categorical, and KL divergence losses. Initially, all losses were relatively high as the VAE worked to reconstruct both the numerical and categorical features. Over time, these losses steadily decreased, indicating the model’s increasing accuracy in capturing the real data distribution. As the training continued, all three losses began to converge, suggesting that the VAE had stabilized and was effectively generating synthetic data that closely aligned with the real patient data.

## Results for fatigue

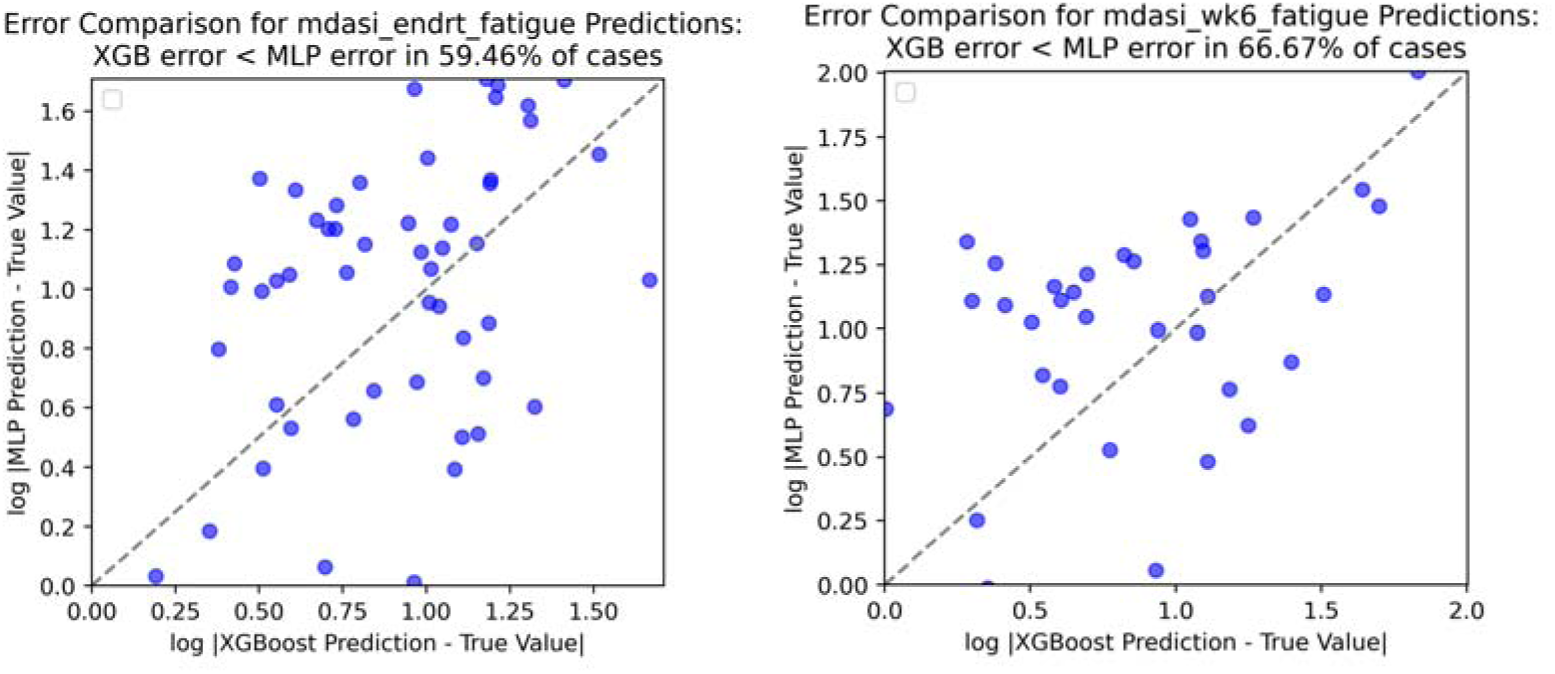
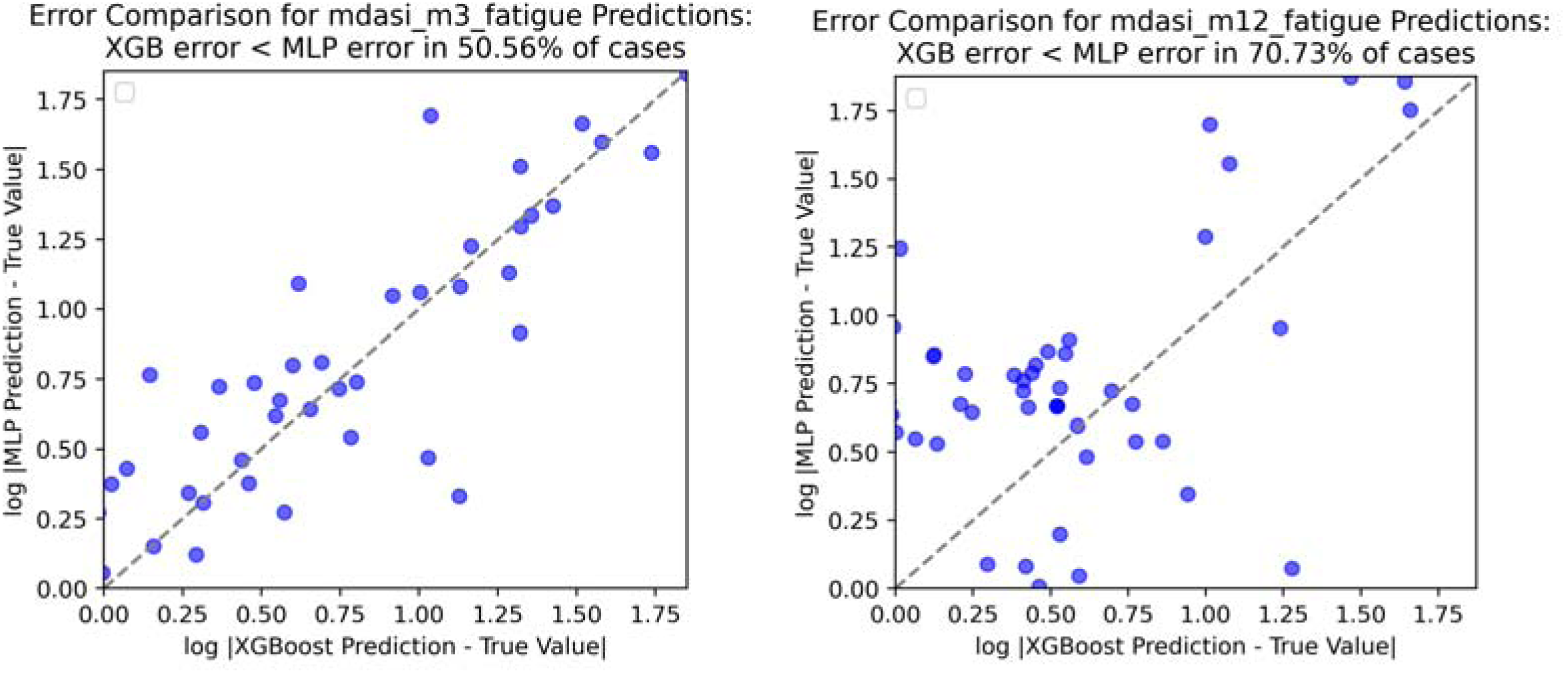

## Results for taste

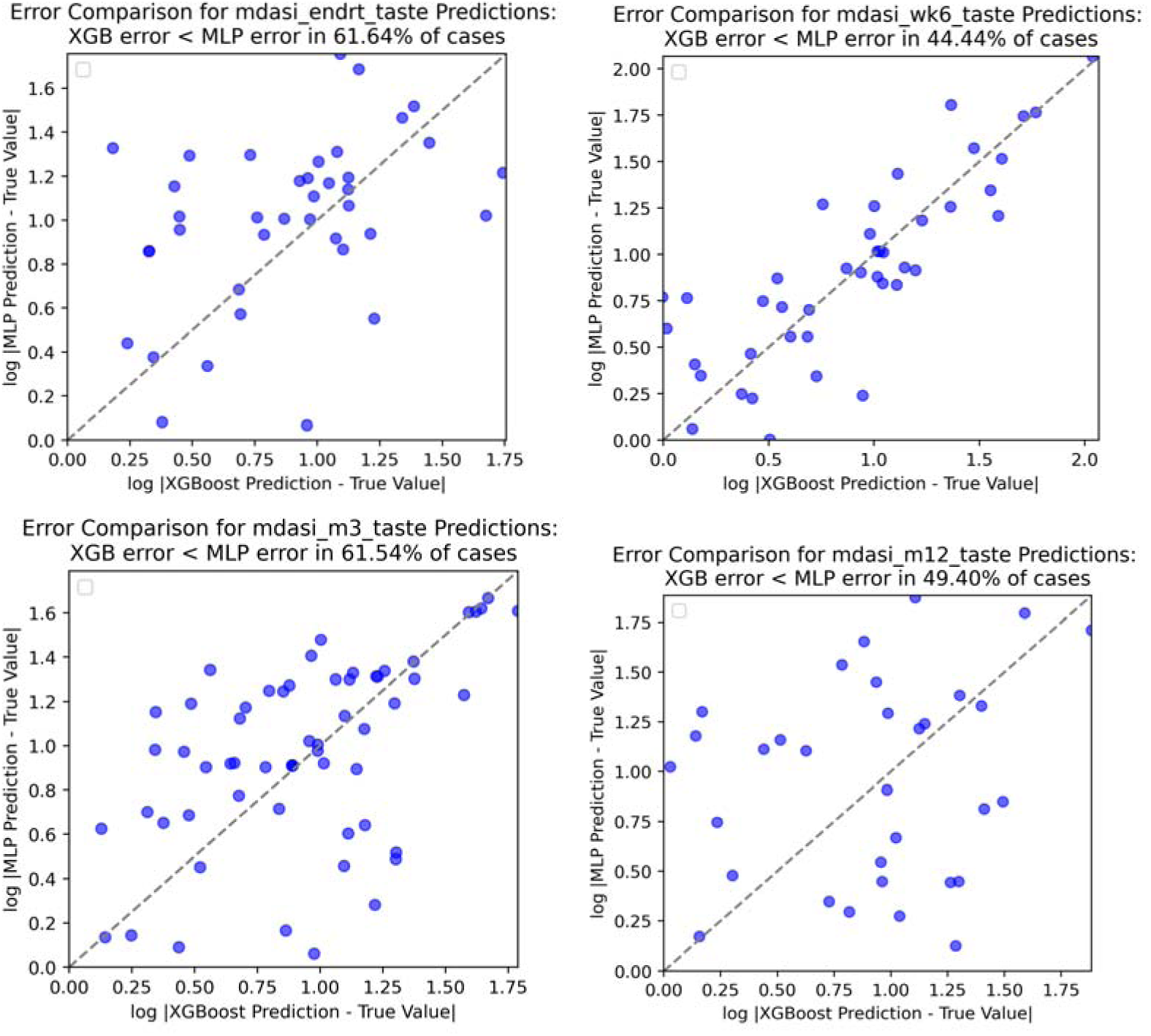

## Results for drymouth

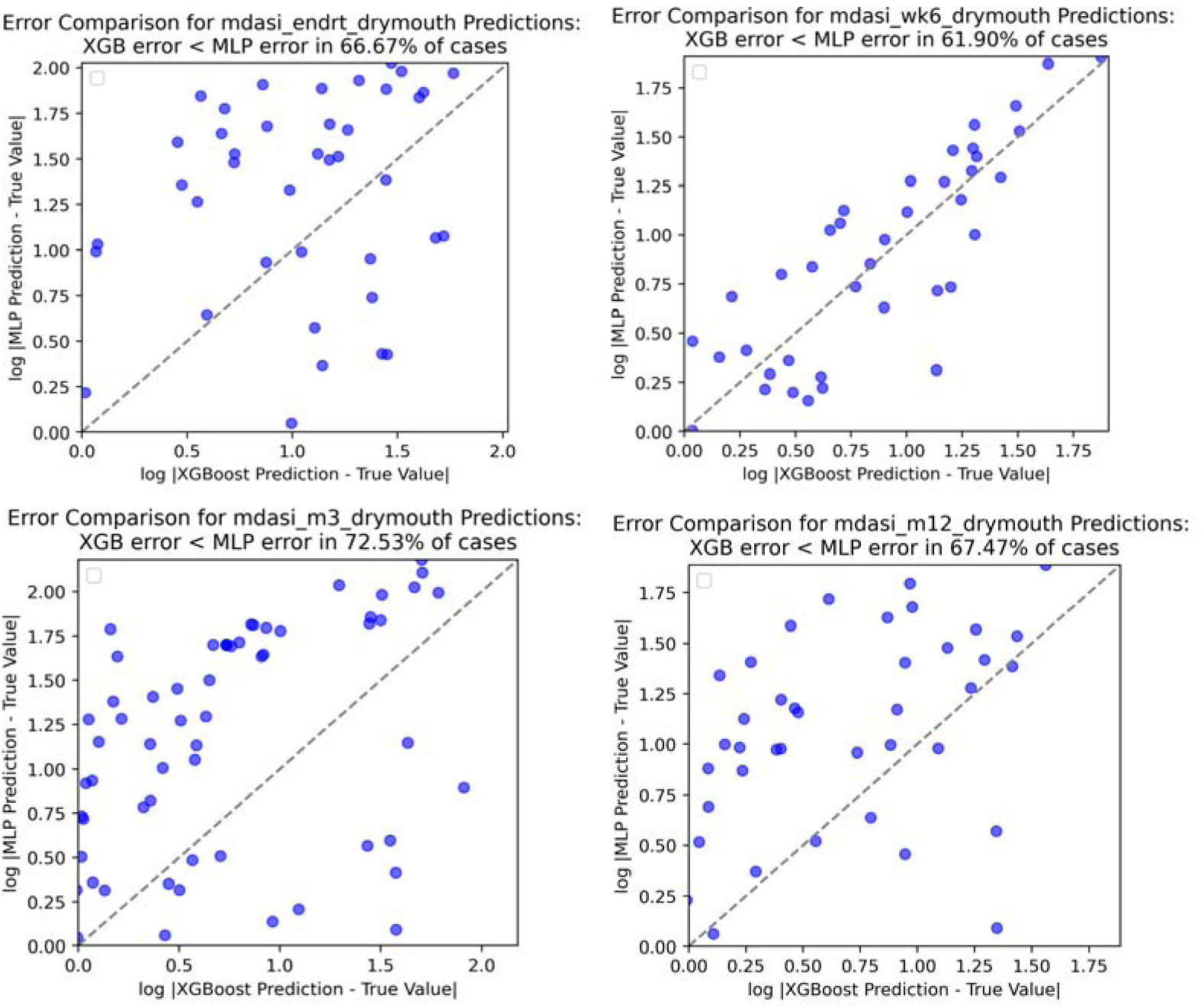

